# Systematic review of candidate prognostic factors for falling in older adults identified from motion analysis of challenging walking tasks

**DOI:** 10.1101/2022.06.23.22275679

**Authors:** Rosemary Dubbeldam, Yu Yuan Lee, Juliana Pennone, Luis Mochizuki, Charlotte Le Mouel

## Abstract

The objective of this systematic review is to identify motion analysis parameters measured during challenging walking tasks which can predict fall risk in the older population. Numerous studies have attempted to predict fall risk from the motion analysis of standing balance or steady walking. However, most falls do not occur during steady gait but occur due to challenging centre of mass displacements or environmental hazards resulting in slipping, tripping or falls on stairs. We conducted a systematic review of motion analysis parameters during stair climbing, perturbed walking and obstacle crossing, predictive of fall risk in healthy older adults. We searched the databases of Pubmed, Scopus and IEEEexplore.

A total of 78 articles were included, of which 62 simply compared a group of younger to a group of older adults. Importantly, the differences found between younger and older adults did not match those found between older adults at higher and lower risk of falls. Two prospective and six retrospective fall history studies were included. The other eight studies compared two groups of older adults with higher or lower risk based on mental or physical performance, functional decline, unsteadiness complaints or task performance. A wide range of parameters were reported, including outcomes related to success, timing, foot and step, centre of mass, force plates, dynamic stability, joints and segments. Due to the large variety in parameter assessment methods, a meta-analysis was not possible. Despite the range of parameters assessed, only a few candidate prognostic factors could be identified: older adults with a retrospective fall history demonstrated a significant larger **step length variability**, larger **step time variability**, and **prolonged anticipatory postural adjustments** in obstacle crossing compared to older adults without a fall history. Older adults who fell during a tripping perturbation had a **larger angular momentum** than those who did not fall. Lastly, in an obstacle course, **reduced gait flexibility** (i.e., change in stepping pattern relative to unobstructed walking) was a prognostic factor for falling in daily life. We provided recommendations for future fall risk assessment in terms of study design.

In conclusion, studies comparing older to younger adults cannot be used to explore relationships between fall risk and motion analysis parameters. Even when comparing two older adult populations, it is necessary to measure fall history to identify fall risk prognostic factors.

## 1 Introduction

Falls in older adults are frequent, with studies in numerous countries reporting fall rates between 15 and 33 % per year for older adults living in the community (Kwan et al., 2011; Peel, 2011). Fall rates increase with age, reaching 50 % for subjects aged more than 85 years (Iinattiniemi et al., 2009) and 60 % for those older than 90 (Fleming et al., 2008). Such falls result in injury in 15 to 45% of the cases (Mackenzie et al., 2002; Kannus et al., 2005), and pose a high economic burden for acute health care and rehabilitation (Heinrich et al., 2010).

A significant amount of research has been aimed at identifying older individuals at increased risk of falling, to orient them to appropriate prevention or rehabilitation programs. These have identified risk factors at the level of individual body functions and structures, such as decreased foot or trunk muscle strength (Mickle et al., 2009; Granacher et al., 2014), cognitive impairments and flexibility impairments (Tinetti et al., 1988; Speechley and Tinetti, 1991). Risk factors have also been identified at the level of task performance, such as walking, Timed-up-and-Go (TUG) or one limb stance (Tinetti et al., 1988; Swanenburg et al., 2010; Ihlen et al., 2018). Machine learning techniques have been used to derive fall risk prediction models, based on multiple candidate prognostic factors (Gietzelt et al., 2014; van Schooten et al., 2015; Ihlen et al., 2016; Greene et al., 2017; Silva et al., 2017; Hua et al., 2018; Rehman et al., 2020). So far, candidate prognostic factors such as step length, step time, cadence and harmonic ratio have been assessed from accelerometer signals recorded in the lab (during gait or TUG) (Greene et al., 2017; Silva et al., 2017; Hua et al., 2018; Rehman et al., 2020; Tunca et al., 2020) or in daily life (10-20 second gait bouts)(Gietzelt et al., 2014; van Schooten et al., 2015; Ihlen et al., 2018). However, the success rate for fall risk prediction varies depending on the locomotion task, with very disparate levels of reported sensitivity (55-100%), specificity (15-100%) and accuracy (62-100%) (Howcroft et al., 2013; Van Schooten et al., 2016; Montesinos et al., 2018; Rehman et al., 2020; Tunca et al., 2020).

The relatively poor performance of these fall risk prediction models may be due to fact that they rely on parameters measured during steady-state locomotion, whereas falling in real life occurs during more challenging locomotion tasks (Tinetti et al., 1988; Speechley and Tinetti, 1991; Luukinen et al., 2000; Mackenzie et al., 2002; Sartini et al., 2010). Indeed, these prospective studies indicate that most falls (60 %) occur during challenging centre of mass (CoM) displacements, such as weight transfers, standing up or sitting down, bending over, or after an external perturbation such as a push or a pull. The next leading cause of falls is the presence of an environmental hazard (30 - 50 %), resulting in slipping, tripping, falls from an upper level (a height) or falls on stairs. Moreover, falls during such challenging locomotion tasks are related to the highest risk of severe injury i.e., fractures (Luukinen et al., 2000).

The objective of our systematic review is to determine which performance parameters assessed during challenging walking tasks are best related to falling in the older adult population. Specifically, we chose to focus on the three biomechanically challenging tasks studied in a laboratory context which are the most representative of falling in daily living: crossing obstacles, ascending and descending stairs, and external perturbations to walking.

## 2 Methods

### 2.1 Literature Search

Factors related to fall risk are ideally studied in a prospective study with older adults. As prospective studies are time-consuming, they are limited in number. Thus, cross-sectional observational studies were also included in this review. Fall risk has been related, among others, to age, to fall history and to physical and mental impairments. Therefore, this review included “ageing studies”, which compare younger adults to older adults and “risk studies”, which compare older adults with a higher fall risk to those with a lower fall risk, determined either prospectively, or based on fall history or mental and physical impairments.

Relevant articles should study the association between fall risk and motion analysis outcome parameters (either kinematic, kinetic, or spatial-temporal parameters). These outcome parameters should be measured during either stair climbing, perturbed walking or obstacle crossing, since these challenging walking tasks are the most related to the circumstances of falls in daily living.

The literature search was performed using the PubMed, Scopus, and IEEExplore search engines. A Boolean combination of the following terms was used to search the aforementioned databases on October 2022: (((fall) OR (fall risk)) AND ((obstacle) OR (stair) OR (perturbation)) AND ((age) OR (older) OR (elderly)) NOT ((diabetes[Title/Abstract]) OR (rheumatoid arthritis[Title/Abstract]) OR (osteoarthritis[Title/Abstract]) OR (Parkinson[Title/Abstract]) OR (stroke[Title/Abstract]))). Furthermore, all articles were published in English and no period restriction was given as a filter. The search string needed to be modified for the Scopus search since initially more than 25 000 articles were returned. Therefore, the search string was modified such that the tasks needed to be reported in the title, the search was restricted to certain domains, certain diseases were excluded if mentioned in title or keywords, and the option to exclude certain study designs and types was used. Details on the search strings used is provided in Appendix A.

For article extraction, two reviewers screened through the titles and abstracts, then the full text. When two reviewers had opposite opinions about the inclusion of an article, a third reviewer made the final decision. The inclusion criteria were: 1) the article examined at least two groups with different fall risk (either younger and older adults, or older adults with higher and lower fall risk); 2) the article reported group differences in kinematic, kinetic or spatiotemporal parameters when performing either stair climbing, perturbed walking or obstacle crossing; and 3) all of the participants were healthy or suffering from only minor impairments corresponding to normal age degeneration, i.e., they should not suffer from any moderate to severe neurological, musculoskeletal disorder, or other conditions related to cognitive disorders and visual impairment. Exclusion criteria were: 1) the article examined the group differences in EMG or EEG signal outcomes; 2) the study compared the difference between a control group and a specific diseased or sensory impaired group, such as diabetes, arthritis, stroke, Parkinson; 3) the study design included an intervention and examined the change after the intervention; 4) dual-task studies. Dual task studies were considered outside the scope of this review, which focused on biomechanically challenging walking tasks. The influence of cognitively challenging tasks on walking performance has been reviewed elsewhere (Smith et al., 2017).

### 2.2 Effect size

The effect sizes reported by the studies were used or we calculated Cohen’s D for each significant finding where effect size was not reported in the article. Cohen’s D was assessed from the deviation of the mean from each group divided by the pooled standard deviation (SD). The equation of Cohen’s D equation is given below (Equation 2):

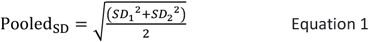

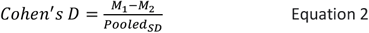

Some studies did not report significant difference levels between groups or between repeated measures. In such cases, the minimum required sample size to reach a significance level of p = 0.05 was calculated in G*power (G*power version 3.1.9.6), based on the reported independent or dependent group means and corresponding standard deviations. If the sample size of the groups was higher than the G*power calculated sample size, we report the finding as a significant difference. Otherwise, we do not report the finding (neither as significant nor as non-significant).

A meta-analysis was not possible, due to the many differences within the experiments as well as in the calculation methods of the outcome parameters in the articles.

### 2.3 Level of evidence

Since multiple types of studies were included, guidelines for systematic reviews of prognostic studies (Riley et al., 2019), observational studies (Mueller et al., 2018; Dekkers et al., 2019), and non-randomised controlled trials (Sterne et al., 2016) were followed. Within these guidelines, required data extraction is similar and includes a description of the study design, participant and sample size, the experiment (a challenging walking related task), analysis method, the outcome measures, and corresponding significant findings (effect estimates).

In the above-mentioned guidelines, bias assessment includes confounding factors and covers selection and information bias, where the signalling questions to determine the bias differ per study type. Bias assessment in prognostic studies can be performed using the bias domains and corresponding signalling questions suggested in QUIPS (Hayden et al., 2013). However, these signalling questions do not cover all selection biases that may occur in observational studies, such as participant group allocation, which is better represented e.g., in the selection bias assessment of the ROBINS-I guideline. For observational studies, however, there is no agreed-upon bias assessment guideline (Mueller et al., 2018; Ma et al., 2020). To ensure the identification of all bias risks in this review, we followed the four crucial steps suggested by (Dekkers et al., 2019) and recommendations made by (Dekkers et al., 2019; Riley et al., 2019). First, a team of reviewers with experience in the field of fall risk, (para-)medical therapy, older adults, machine learning, and systematic reviews was initiated. Second, our *target trial (gold standard)* was defined as a prospective observational study of older adults including an assessment of a challenging walking task (experiment/observation) followed by a long term and repeated evaluation of the occurrence of a fall (event). Related to the research question of this review, the aim of the target trial would be to study the relationship between the occurrence of a fall and the task performance outcome measures. The assumption would be that motion analysis outcome parameters with an observed strong relationship with fall occurrence, are candidate prognostic factors for the event of a fall. Third, the effect of interest is defined as the allocation of participants to a group representing fallers (high fall risk) or non-fallers (low fall risk) and how this may influence or bias the outcome parameters.

In the fourth step, the confounding factors and bias domains were discussed and determined, and corresponding signalling questions were defined. In total, seven bias domains were defined, and they relate to potential bias issues occurring before (domain 1, 2), during (domain 3) and after (domain 4, 5, 6, 7) the effect of interest, i.e., allocation of the participants to the fall (risk) group. The first three bias domains include bias distinct from the target trial, such as bias due to confounding, bias due to selection of participants, and bias in the assessment and classification of fall risk. The confounding factors are related to both fall risk group assessment and outcome parameter: i.e., age, gender, mental and physical fitness, frailty, and fall history. Selection bias occurs when participants do not adequately represent the target population. Bias in classification occurs when participants are allocated to the wrong fall risk group, e.g., due to errors in recall or non-valid fall risk assessment methods. For articles comparing older adults at high and low risk of falling, if fall risk was assessed based on fall history, this was considered as a low risk of bias. If fall risk was assessed in another way (typically clinical tests or questionnaires), this was considered a moderate or serious risk of bias, depending on the method used. For articles comparing younger and older adults, the classification bias was considered “not applicable”, and the results from these articles are presented separately. Selection and classification bias only refers to factors related to fall risk (internal validity), not to factors related to generalizability or applicability of the study (external validity).

The other 4 bias domains are independent of the study type and refer to the observation, i.e., the experiment and data handling, and include: bias due to deviations from the intended experiment, bias in the measurement of outcome parameters, bias due to missing data, and bias in the selection of the reported result. Bias due to deviations from the intended experiment may occur if fatigue differentially affects the performance the different groups (for example, older or frailer subjects may be more fatigued towards the end of the experiment than younger or healthier subjects). The motion analysis outcome parameters may be biased if assessors are aware of group status, if different methods are used to assess outcomes in the different groups or if measurement errors are related to group status. Some of the biases from domains 4 (intended experiment) and 5 (outcome parameters) may typically be avoided using blinding. Regarding missing data, enough data should be presented in both groups to be confident of the findings and the missing data should not be group dependent. Bias in reporting the results can occur when the studies only report group means and standard deviations, but not significance level.

To make the scoring repeatable, signalling questions, corresponding sub-questions and bias examples were used (Appendix B, Table B.1). The questions were answered with: ‘no’, ‘probably not’, ‘yes’ or ‘probably yes’. If (probably) no bias was assumed for the signalling question, we moved on to the next signalling question. If bias was assumed, for some domains, corresponding sub-questions were answered. If the signalling and sub-question could not be answered due to the lack of information in the article, the question was scored as ‘no information’. Lastly, for each included article and each bias domain, each bias issue was described, scored qualitatively (low, moderate, serious, no information). The bias scoring into low, moderate, or serious bias was followed as described in QUIPS and ROBINS-I, and detailed in Appendix B, Table B.2. Bias levels were discussed and decided upon, keeping in mind to which extent and in which direction a bias factor might influence the estimated effect compared to the true effect (where the effect is the difference in outcome parameters between groups). The complete risk of bias assessment for all reviewed studies is provided in Appendix C.

In summary, for each included article the study design, included population and sample size, the experiment, the analysis method and the motion analysis outcome parameters (including both significant and non-significant results) were reported. Then, the seven bias domains were evaluated as described above.

## 3 Results

### 3.1 Overview of the selected articles

#### 3.1.1 Article extraction

An overview of the systematic article extraction is given in Figure 1. In total, 2269 articles were extracted from the three databases. First, 376 duplicates were removed. Another 1790 articles were removed based on their titles and abstracts. After reading the full text, further 25 articles were removed for the following reasons: 23 articles focused on dual-tasks, static balance, single steps or steady walking, one article was a systematic review, and one article lacked the description of the participants. In the end, 78 articles were included in this review.

**Figure 1:**
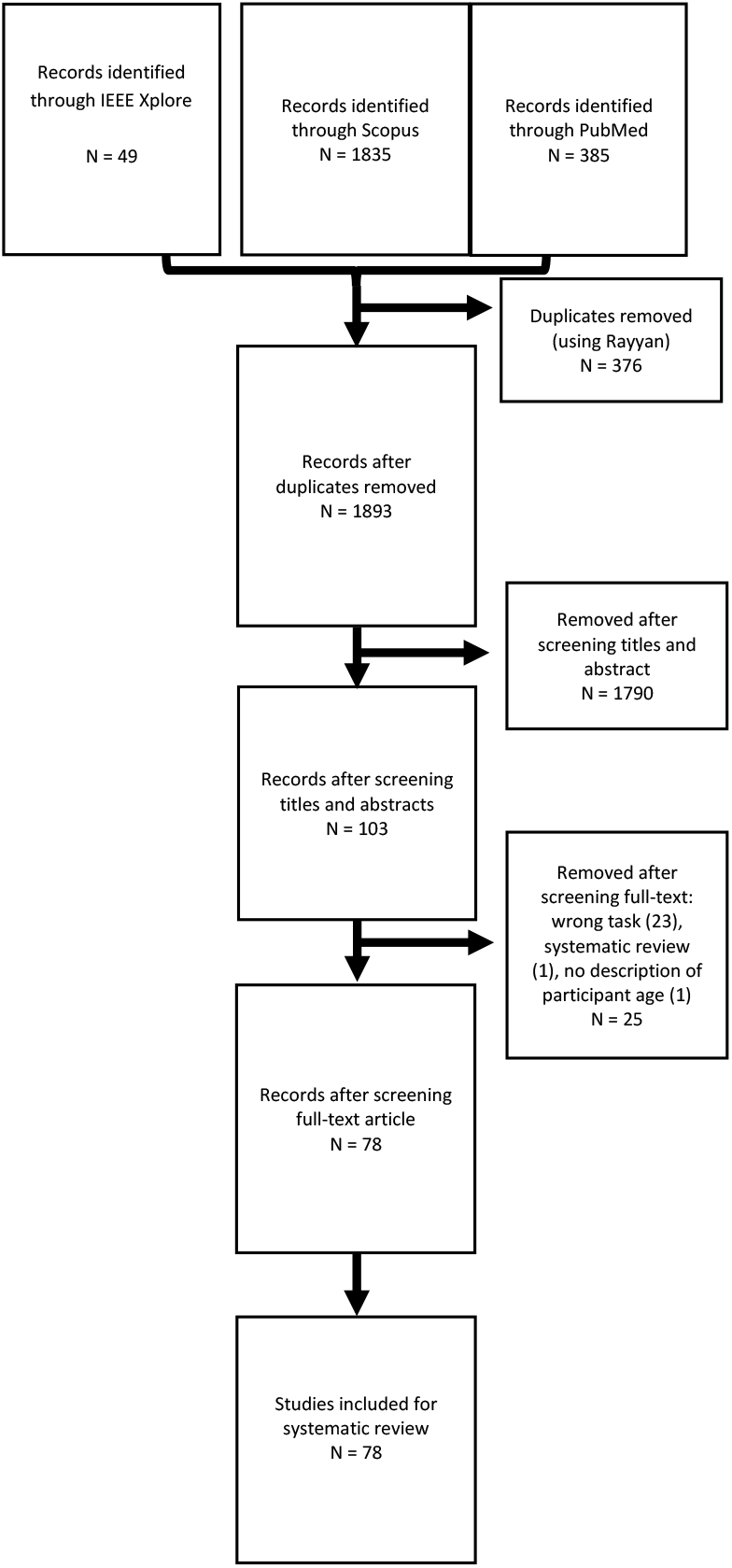
Overview of the systematic article extraction

#### 3.1.2 Fall risk evaluation

Sixteen studies compared a group of higher-risk older adults (mean age ranging from 62.5 to 81.6 years) to a group of lower-risk older adults (mean age ranging from 65.6 to 80.8 years). These studies will be referred to as **risk studies**. The details of the study designs and populations of risk studies are reported in Table 1. Two older adult performance studies assessed fall risk prospectively (Ackermans et al., 2021; Hansson et al., 2021), by following subjects for one year to determine whether they fall, after they performed the challenging walking task. Six studies assessed fall risk retrospectively (Brach et al., 2011; Oh-Park et al., 2011; Uemura et al., 2011; Ackermans et al., 2019; Pieruccini-Faria and Montero-Odasso, 2019; Guadagnin et al., 2020; Gerards et al., 2021), by asking subjects at the time of the walking measurement whether they had fallen in the previous months. Three studies evaluated risk based on physical or mental performance at the time of the walking measurement using clinical tests or questionnaires (Zietz et al., 2011; de Carli et al., 2014; Pan et al., 2016). Two studies evaluated risk based on whether the subjects experienced functional decline or improvement over a one-year follow-up (Brach et al., 2011; Oh-Park et al., 2012). One study divided the subjects into higher and lower risk depending on whether they fell during the challenging walking task itself (Pijnappels et al., 2005). One study compared patients with complaints of “unsteadiness” during walking (higher risk) to a group of healthy controls without a history of falls (Chou et al., 2003). The final study compared a group of hospitalised subjects (higher risk) with a group of healthy subjects (Brodowski et al., 2021). Within each study, the two groups were typically age matched, except for two studies (Zietz et al., 2011; Brodowski et al., 2021). In those two studies, the older adults at higher risk were significantly older than those at lower risk, and this was considered a serious risk of confounding bias (Appendix C, Table C.1).

**Table 1.**
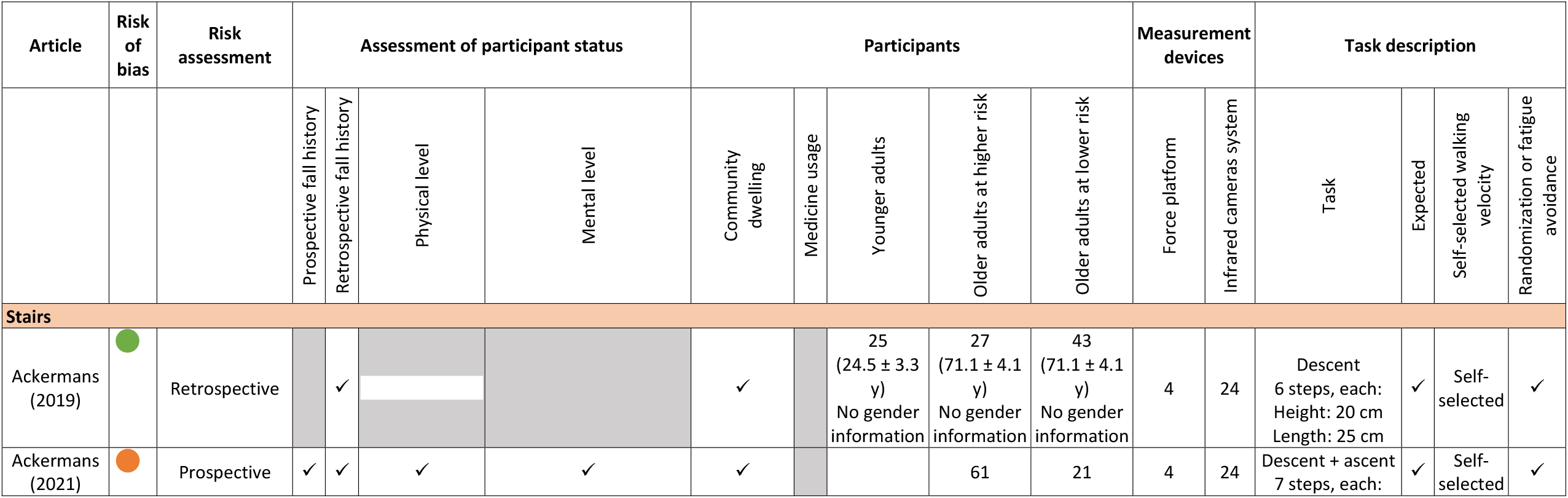
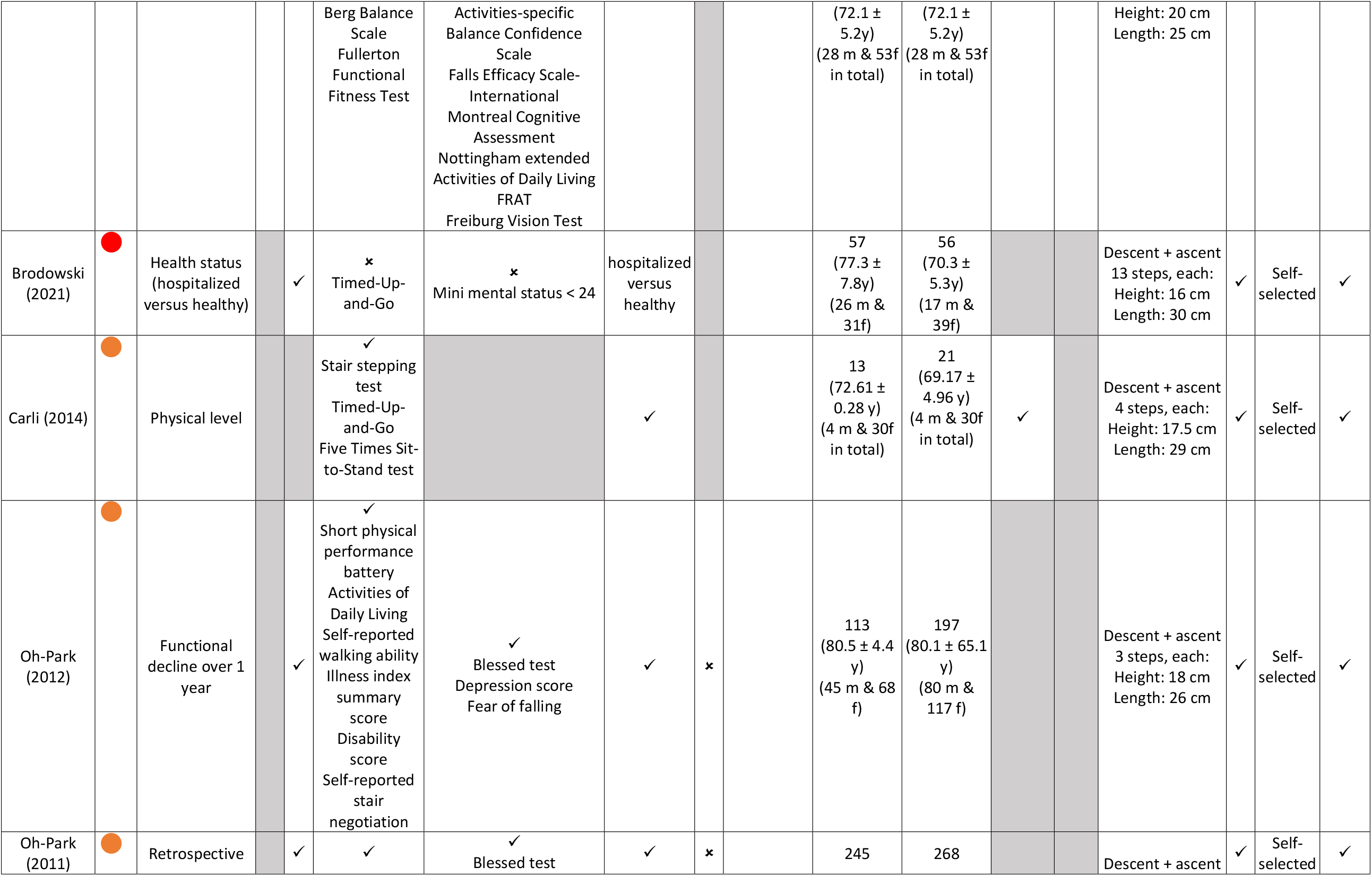
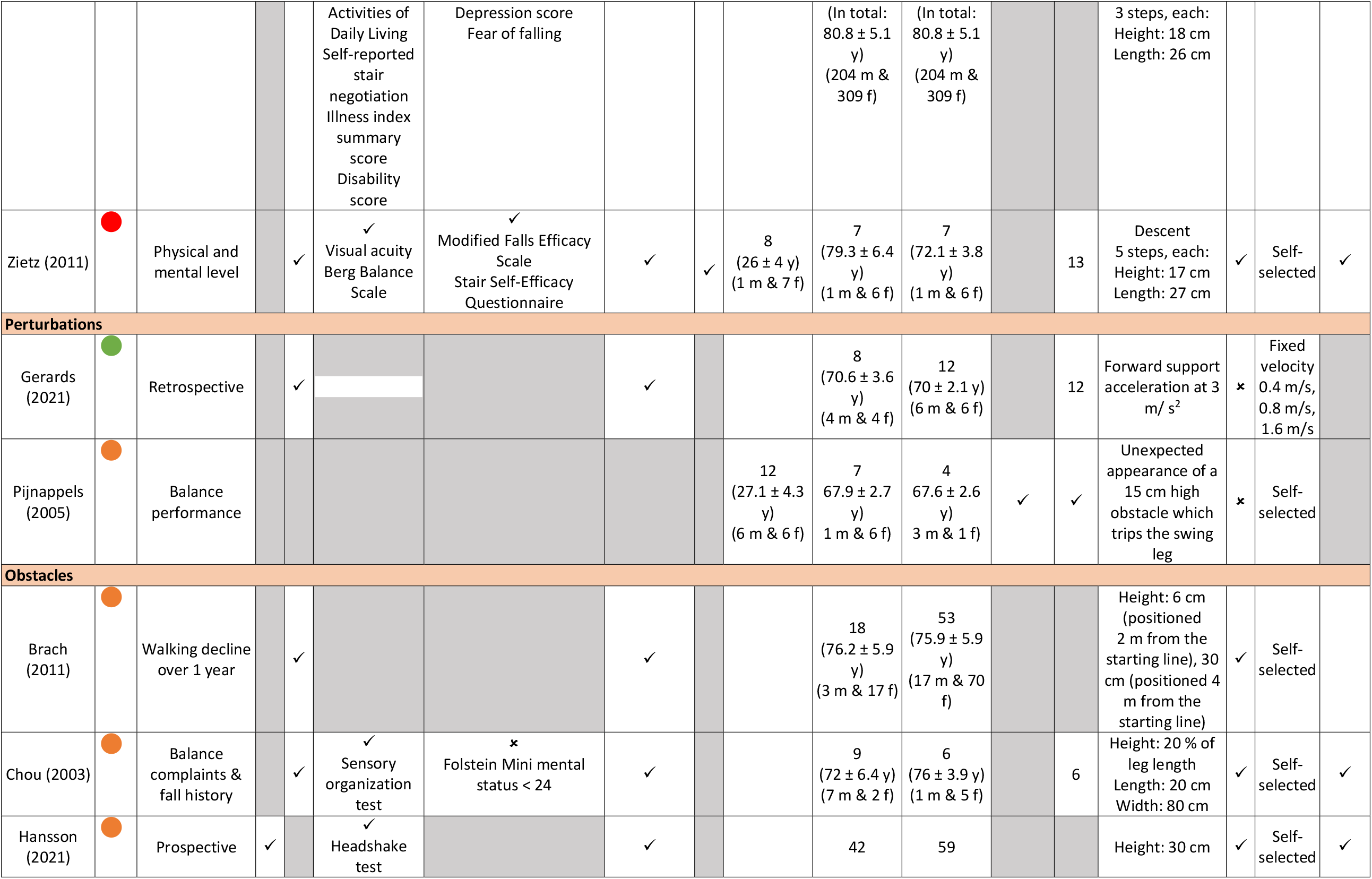
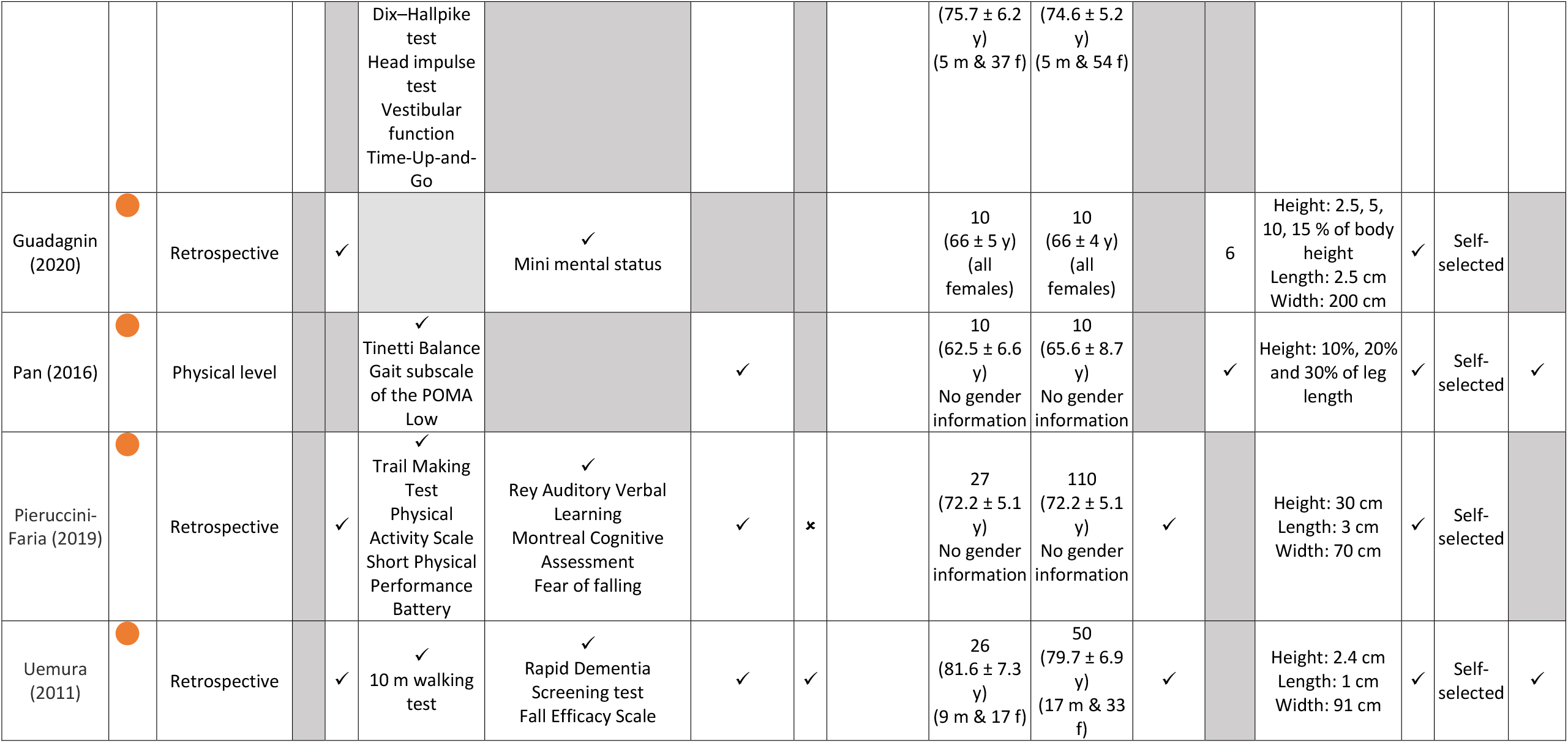
Description of the risk studies, ordered by gait task. Abbreviations: years old: y; male: m; female: f. Shading: 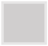 not reported Risk of bias: 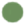 Low, 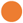 Moderate, 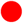 Serious Risk assessment: • Prospective/Retrospective: comparison of older adults who did and did not fall during a period following/preceding the measurement Assessment of participant status: • Prospective fall history: ✓ if the subjects were prospectively followed-up after the measurement to determine whether they fall • Retrospective fall history: ✓ if the subjects were asked whether they had fallen in the months preceding the measurement • Physical/mental level: ∘ ✓ if physical/mental level was tested ∘ ✗ if physical/mental level was tested and subjects with low fitness were excluded from the study. Medicine usage • ✓ if participants took medication during the measurement period • ✗ if participants did not take medication during the measurement period Measurement devices: if force platforms or infrared cameras were used, but their number was not reported, this is indicated by ✓ Task description: Expected: ∘ ✗ if unexpected changes in the task occurred from trial to trail (for example: if various obstacles or stairs were used and these were presented in random order, or if perturbations occurred at random times) ∘ ✓ otherwise

Sixty-two of the selected studies evaluated fall risk based solely on age, comparing a group of younger (mean age ranging from 20.9 to 29.3 years) and a group of older participants (mean age ranging from 55.6 to 81 years). These studies will be referred to as **ageing studies** in the rest of the text. The details of the study designs and populations of ageing studies are reported in Appendix D. The full list of references for ageing studies is in Appendix E.

#### 3.1.3 Tasks

Task characteristics are also reported in Table 1 for risk studies and Appendix D for ageing studies.

Stair climbing was assessed in 7 risk studies and 19 ageing studies. The stairs had a variety of configurations, ranging from a single (Begg and Sparrow, 2000; Crosbie and Gan, 2003) to 13 steps (Brodowski et al., 2021) and the studies evaluated either ascent, descent or both.

Perturbed walking was assessed in 2 risk studies and 20 ageing studies. Most perturbations were applied through a translation of the support surface (14 studies), either in the mediolateral or anteroposterior direction, or both. The other types of perturbations were waist-pulls (Laudani et al., 2021; Rum et al., 2021), ankle pull (McCrum et al., 2016; Bosquée et al., 2021), tripping (Pijnappels et al., 2005), visual perturbations (Sun et al., 2017; Kazanski et al., 2020), soapy water (Liu and Lockhart, 2009) and surface drop (Jeon et al., 2022).

Obstacle crossing was studied in 7 risk and 23 ageing studies. The obstacles used had a variety of dimensions, with a height ranging from 0 cm (visually projected obstacles in (Caetano et al., 2016) and (Chen et al., 1994) up to 30 cm (Pieruccini-Faria and Montero-Odasso, 2019; Hansson et al., 2021) or 30% of the leg length (Lu et al., 2006; Huang et al., 2008; Oh-Park et al., 2012; Park and Lee, 2012; Pan et al., 2016). In the prospective study by Hansson et al., the participants navigated an obstacle course comprising several tasks in sequence: standing up from a chair, walking along a narrow path of 25 cm width for 3 meters, walking over an uneven surface, crossing 3 obstacles of 30 cm height, and climbing up and down a stair of 10 steps (Hansson 2021).

In 62 out of 78 studies, subjects were allowed to walk at their preferred velocity.

Most studies used either an infrared camera system (33 studies), force platforms (10 studies), or both (28 studies) to measure walking parameters. One study used an inertial measurement unit (IMU) attached to the right thigh to measure kinematic parameters (Hansson et al., 2021).

#### 3.1.4 Sample size

The selected studies had a wide range of sample sizes, with 30 studies having 10 or fewer participants in one of the groups, and 2 studies having more than 370 participants in total (Oh-Park et al., 2011, 2012). Overall, risk studies assessed 680 older participants with a higher risk and 927 with a lower risk. Ageing studies assessed 921 younger participants and 1065 older participants.

#### 3.1.5 Level of evidence

An overview of the risk of bias for each domain and article is given in Appendix C. Based on the seven risk of bias domains, we classified 23 studies with low risk of bias, 48 studies with moderate risk of bias and 7 studies with serious risk of bias. Typical biases among the articles included in the seven domains were:

1. Confounding factors: confounding due to a difference in the gender ratio between groups (9 studies); no information on the gender ratio between groups (18 studies).
2. Participant selection: the health status of the participants (physical health, mental health or fall history) was used an exclusion criterion (15 studies); the study population had an unbalanced gender ratio (32 studies, including 11 which included either only females or only males).
3. Group allocation: risk was evaluated based on balance tests or clinical tests or questionnaires rather than prospective or retrospective fall history (6 studies)
4. Intended experiment: no information on either randomization or fatigue prevention (42 studies)
5. Outcome parameters: invalid assessment of centre of mass location (3 studies)
6. Missing data: missing data due to differences or errors in task performance resulting in unbalanced groups for analysis (10 studies).
7. Result reporting: significance level was not reported (4 studies).

### 3.2 Motion analysis outcome parameters

A wide range of parameters were reported by the studies, including outcomes related to success, timing, foot and step, centre of mass, force plates, dynamic stability, joints and segments. Table 2 presents the subset of outcome parameters that were reported for at least one risk study, and indicates the studies reporting either significant or non-significant findings for each outcome. Outcome parameters that were only reported in ageing studies are listed in Appendix F. The most commonly reported parameters were step length (7 risk and 31 ageing studies), stance, swing and compensatory duration (6 risk and 23 ageing studies) and walking, approaching or crossing speed (11 risk and 20 ageing studies). Thirty-seven parameters were reported by a single study in a single task.

**Table 2:**
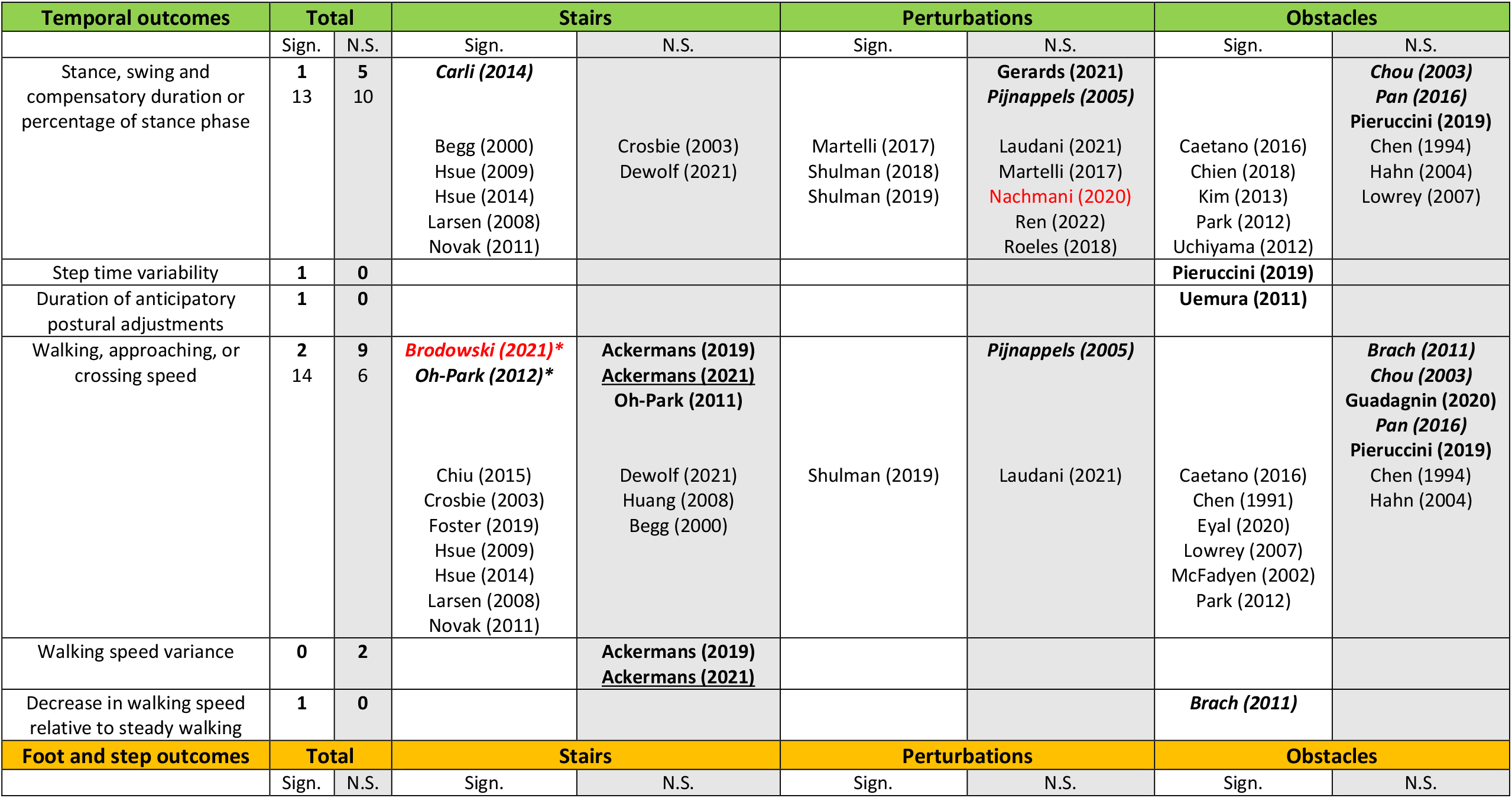
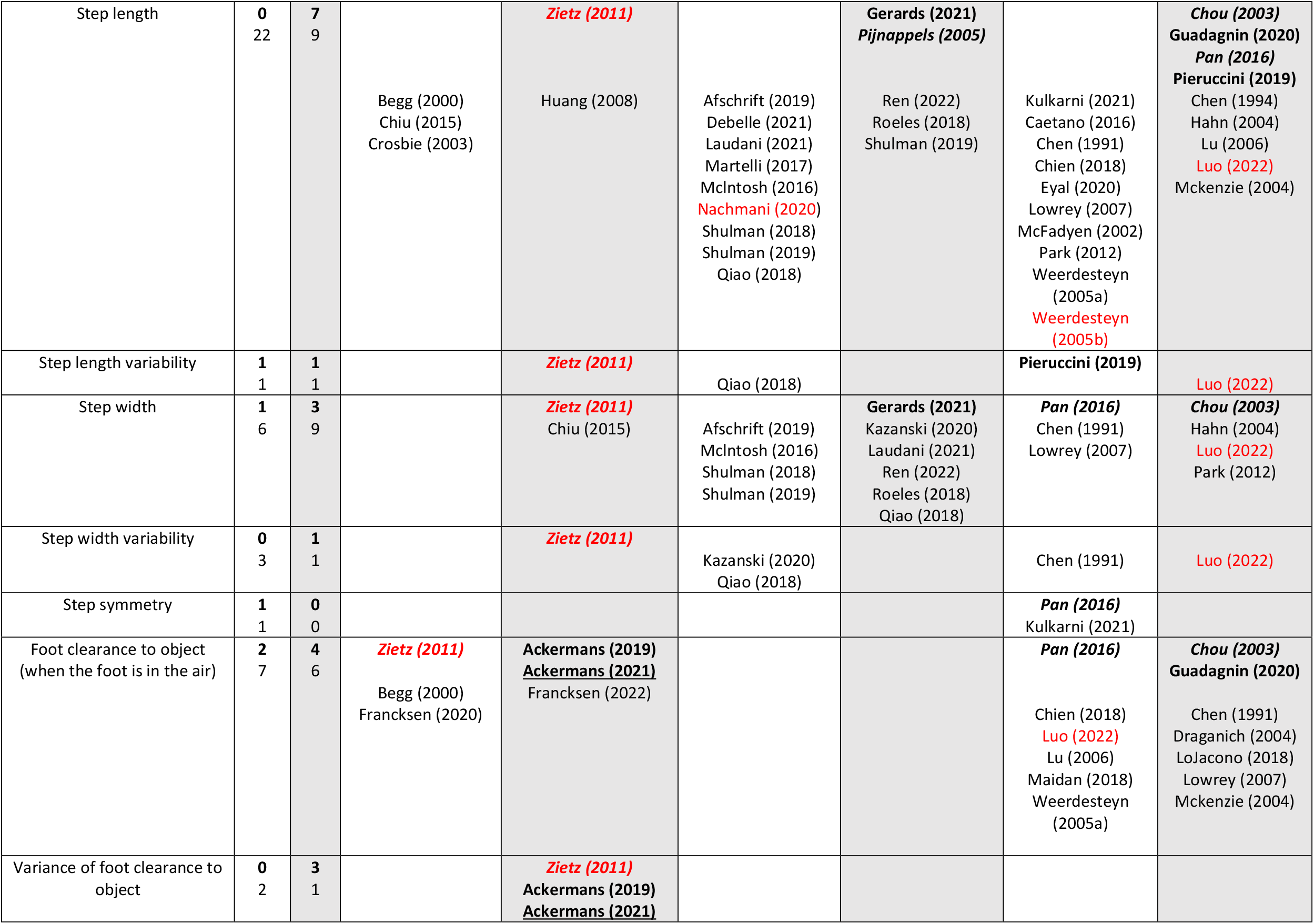
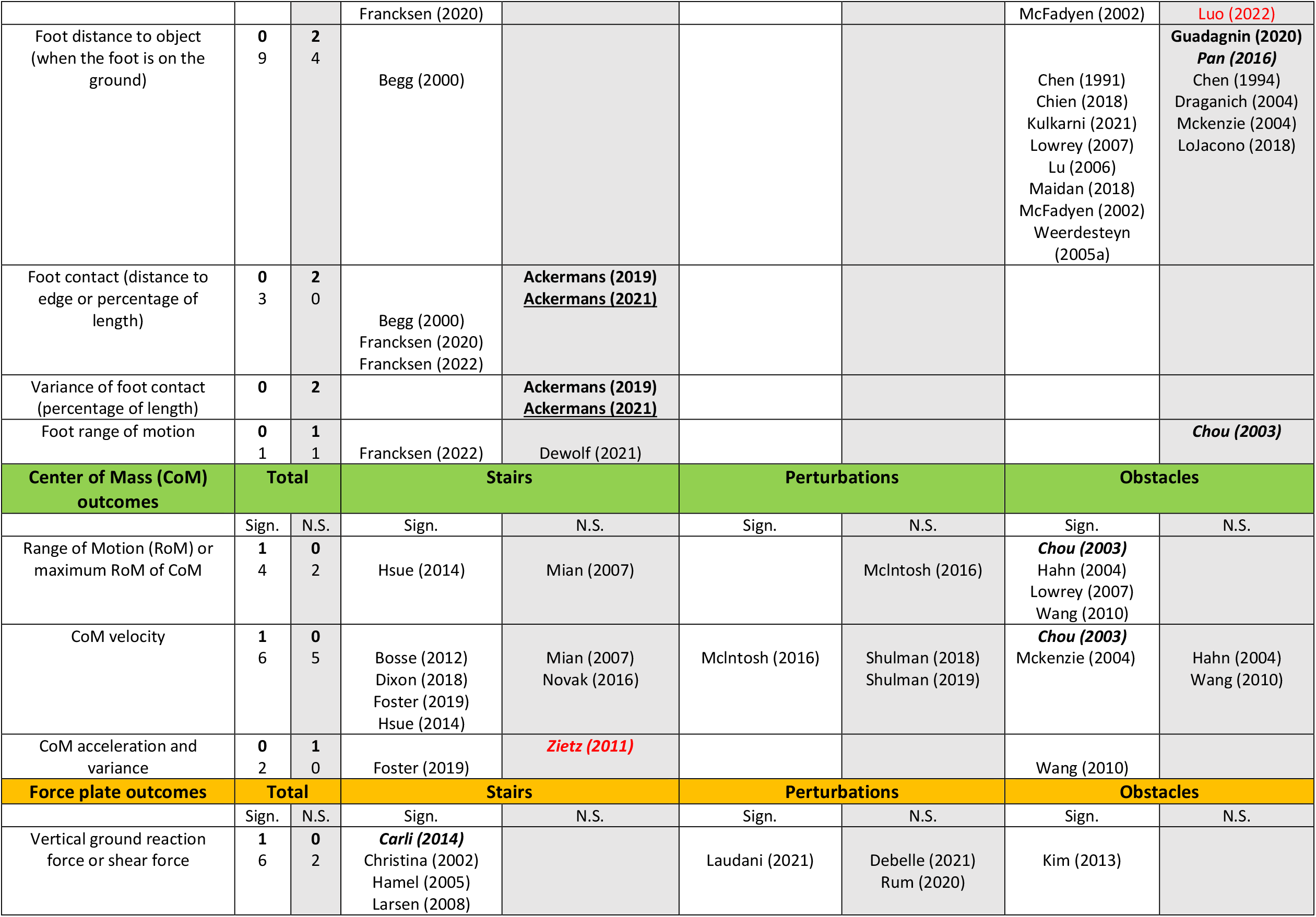
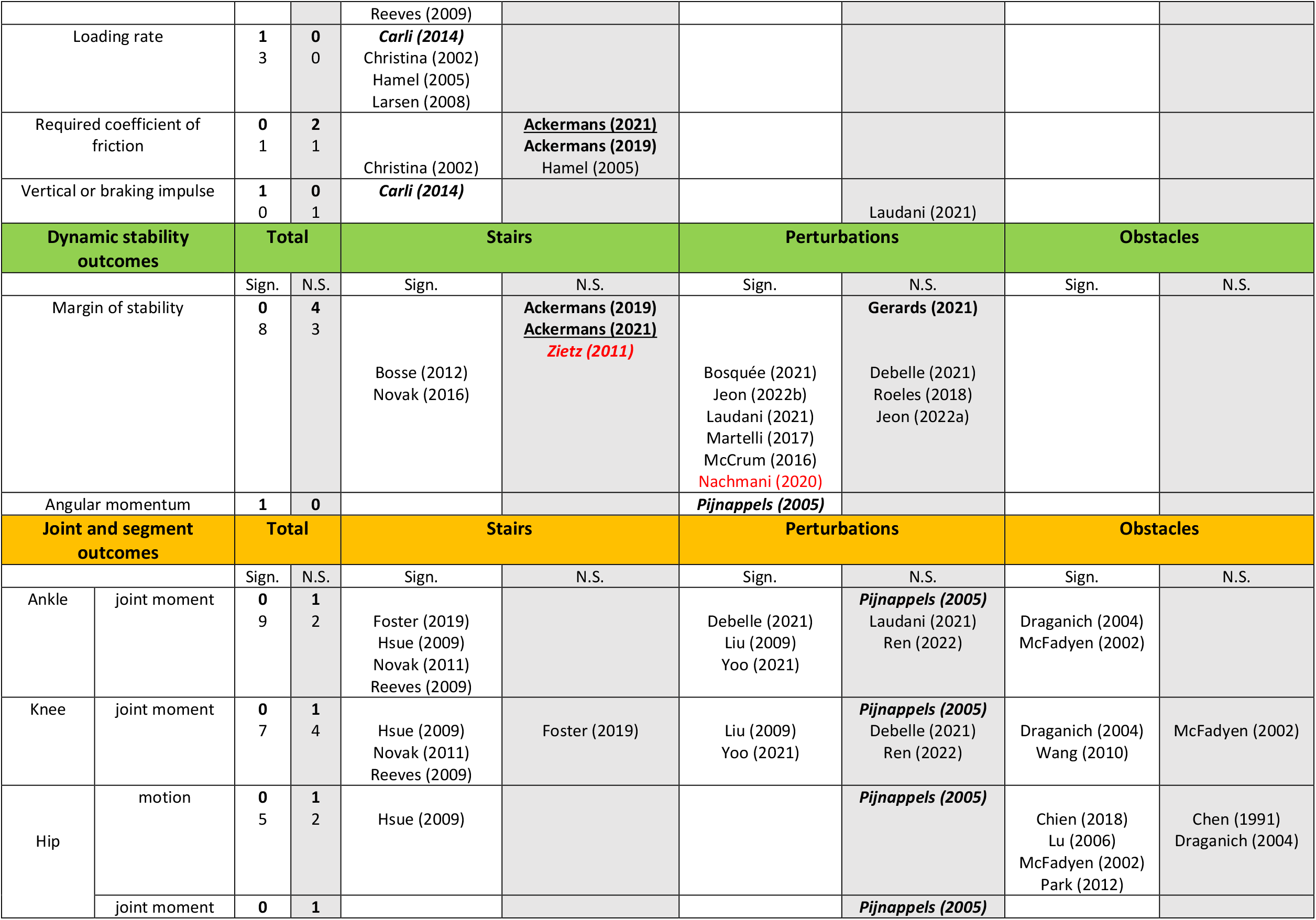
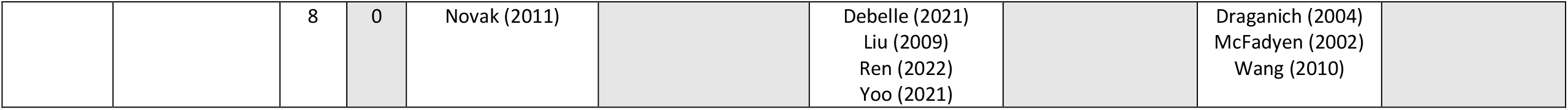
Significant and non-significant findings in risk studies Overview of the reported outcome parameters and corresponding articles reporting either significant or non-significant findings. **Risk studies** (comparing older adults with higher and lower risk) are in bold, and underlined if fall risk is assessed **<u>prospectively</u>**, or in italics if risk is evaluated based on ***physical or mental level***. Ageing studies (comparing younger and older adults) are not in bold. Studies with a serious risk of bias are in red. Studies for which the significance level was not reported, but for which the sample size was sufficient (see G*method calculation in the Methods) are indicated with an asterisk (*). Abbreviations: Sign. significant; N.S. non-significant.

#### 3.2.1 Lack of agreement between ageing and risk studies

An important finding is that outcomes which were significantly different between younger and older adults were not necessarily good prognostic factors for fall risk (Table 2). This finding was particularly robust when considering outcomes reported by a large number of studies.

The most reported finding was step length, which was found to be significantly shorter between younger and older adults in a majority of studies (Table 2) for stairs (3 out of 4 studies), perturbations (9 out of 12 studies) and obstacles (10 out of 15 studies). In contrast, step length was not significantly different between older adults at higher and lower risk in either stairs (Zietz et al., 2011), perturbations (Pijnappels et al., 2005; Gerards et al., 2021) or obstacles (Chou et al., 2003; Pan et al., 2016; Guadagnin et al., 2020; Gerards et al., 2021). This finding was consistent whether risk was evaluated based on retrospective fall history (Pieruccini-Faria and Montero-Odasso, 2019; Guadagnin et al., 2020; Gerards et al., 2021), balance performance (Pijnappels et al., 2005), physical and mental level (Zietz et al., 2011; Pan et al., 2016) or unsteadiness complaints (Chou et al., 2003).

Another commonly reported finding was speed when walking over obstacles. The speed was found to be significantly higher in younger than older subjects in 8 out of the 10 studies which assessed this parameter (Table 2). In contrast, no significant difference in obstacle walking speed between older adults at higher and lower risk was found in any of the 5 studies that assessed this parameter, whether risk was evaluated based on retrospective fall history (Pieruccini-Faria and Montero-Odasso, 2019; Guadagnin et al., 2020), physical level (Pan et al., 2016), unsteadiness complaints (Chou et al., 2003) or walking decline over a year (Brach et al., 2011).

Since the goal of this review was to identify candidate fall risk prognostic factors for older subjects, in the rest of this result section we only report the results from risk studies.

#### 3.2.2 Walking, approaching, and crossing speed

In most studies (62 out of 78), subjects were allowed to walk at self-selected speed. Walking, approaching or crossing speed was assessed in 5 risk studies with stairs, 1 risk study with perturbations and 5 risk studies with obstacles. Stair studies reported speed in number of steps per second (Brach et al., 2011; Ackermans et al., 2019, 2021) and two studies reported the total stair ascent or descent time (Oh-Park et al., 2011, 2012). The perturbation study reported speed in meters per second (Pijnappels et al., 2005). Three obstacle studies used multiple steps (including the obstacle crossing steps) to assess an average walking speed, either over several meters or over 6 steps (Brach et al., 2011; Pieruccini-Faria and Montero-Odasso, 2019; Guadagnin et al., 2020). One obstacle study reported the crossing speed of the single stride over the obstacle (Pan et al., 2016).

As mentioned previously, none of the obstacle studies found a significant difference in speed between older adults at higher and lower risk (Figure 2). While subjects significantly slowed down when crossing obstacles with increasing height, this did not differ significantly between fall risk groups (Pan et al., 2016). One study reported the decrease in walking speed when walking over an obstacle compared to normal walking (Brach et al., 2011). Whereas the walking speed over the obstacle itself was not significantly different across groups, the decrease in speed compared to baseline was significantly larger for older subjects whose walking speed declined over a one-year follow-up (Brach et al., 2011). Note that in this study, retrospective fall history was also assessed, and was not significantly different between subjects whose walking speed improved or declined over one year.

**Figure 2:**
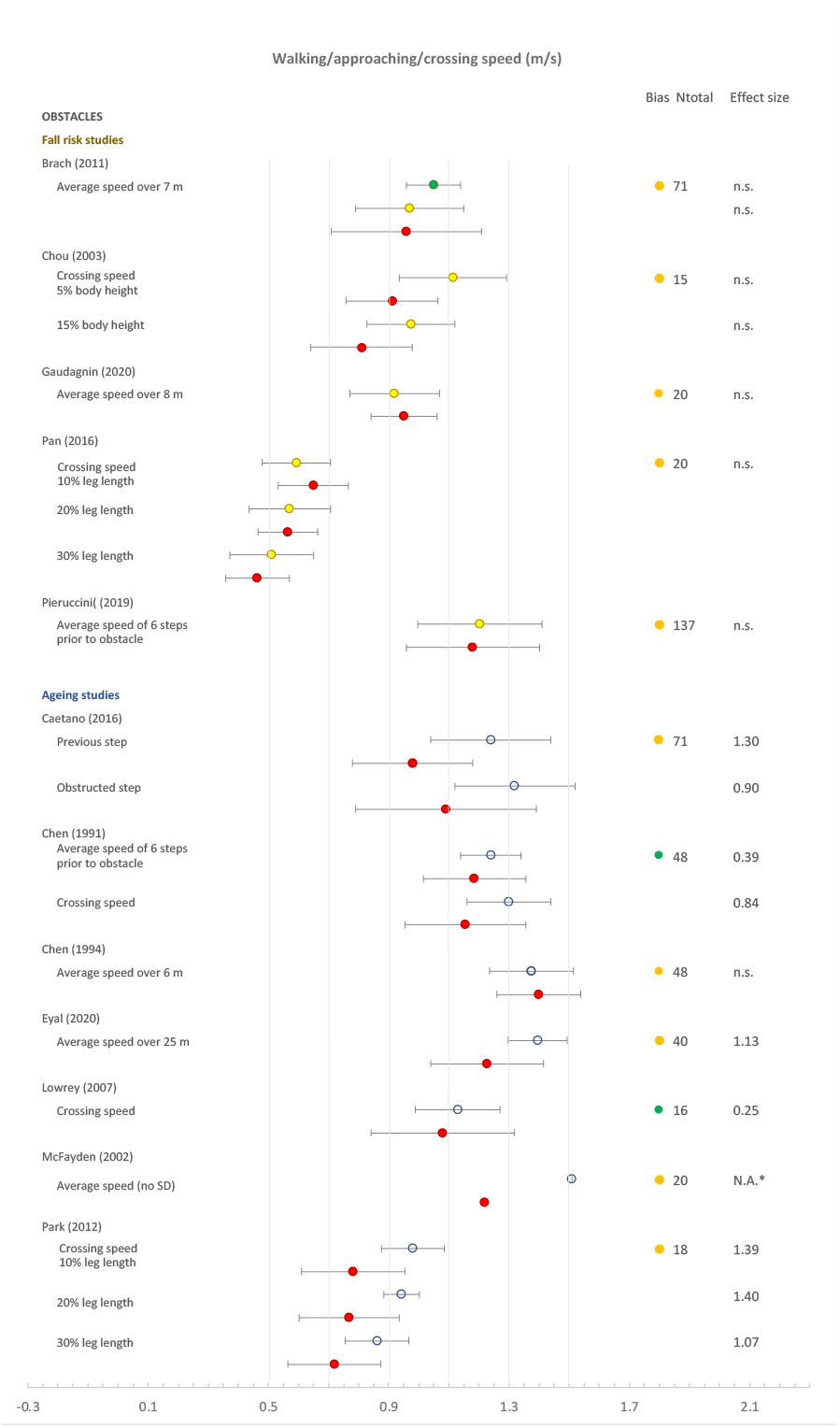
Forest plot of walking speed. Circles: open blue– younger adults, yellow closed– older adults with low fall risk, red closed – older adults with high fall risk, brown closed– older adults with unspecified fall risk. For Brach (2011): older adults whose gait speed improved / stayed the same / deteriorated after a year are indicated in green / yellow / red.; m/s, meters per second; N.A.*, not applicable, the standard deviation was not given; n.s., not significant. Ntot is the total number of participants.

**Figure 3:**
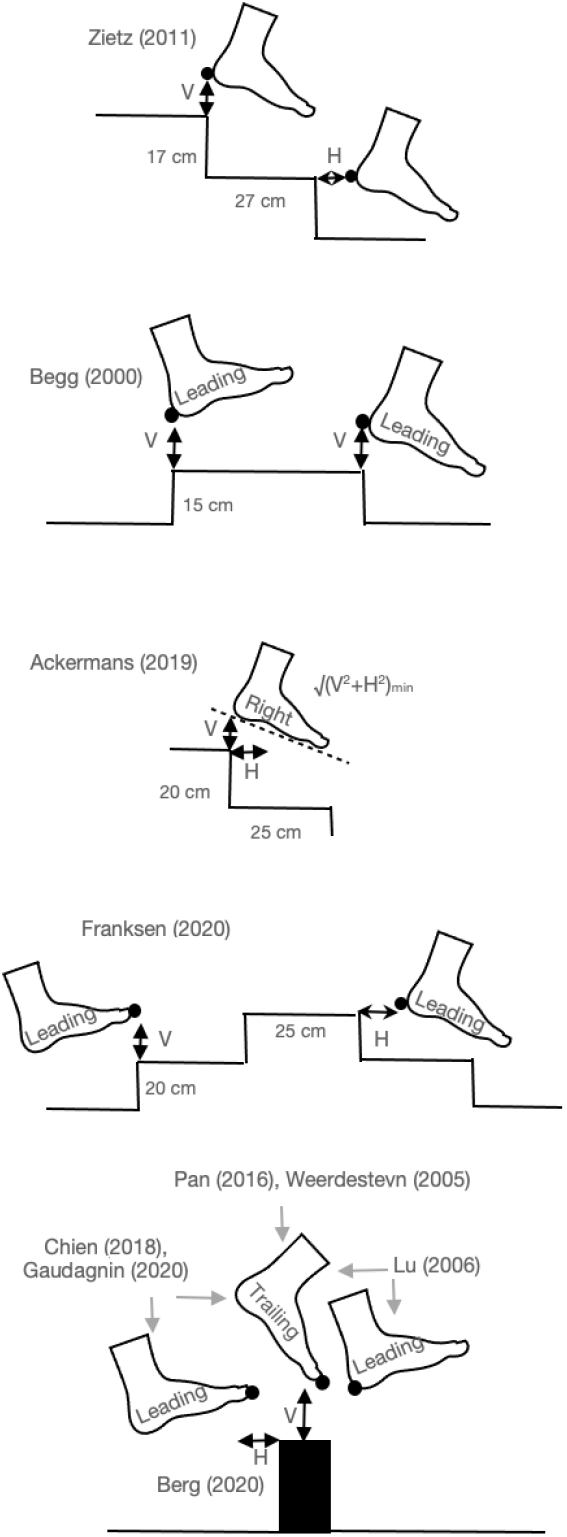
Foot clearance definitions from different studies. V, vertical; H, horizontal.

**Figure 4:**
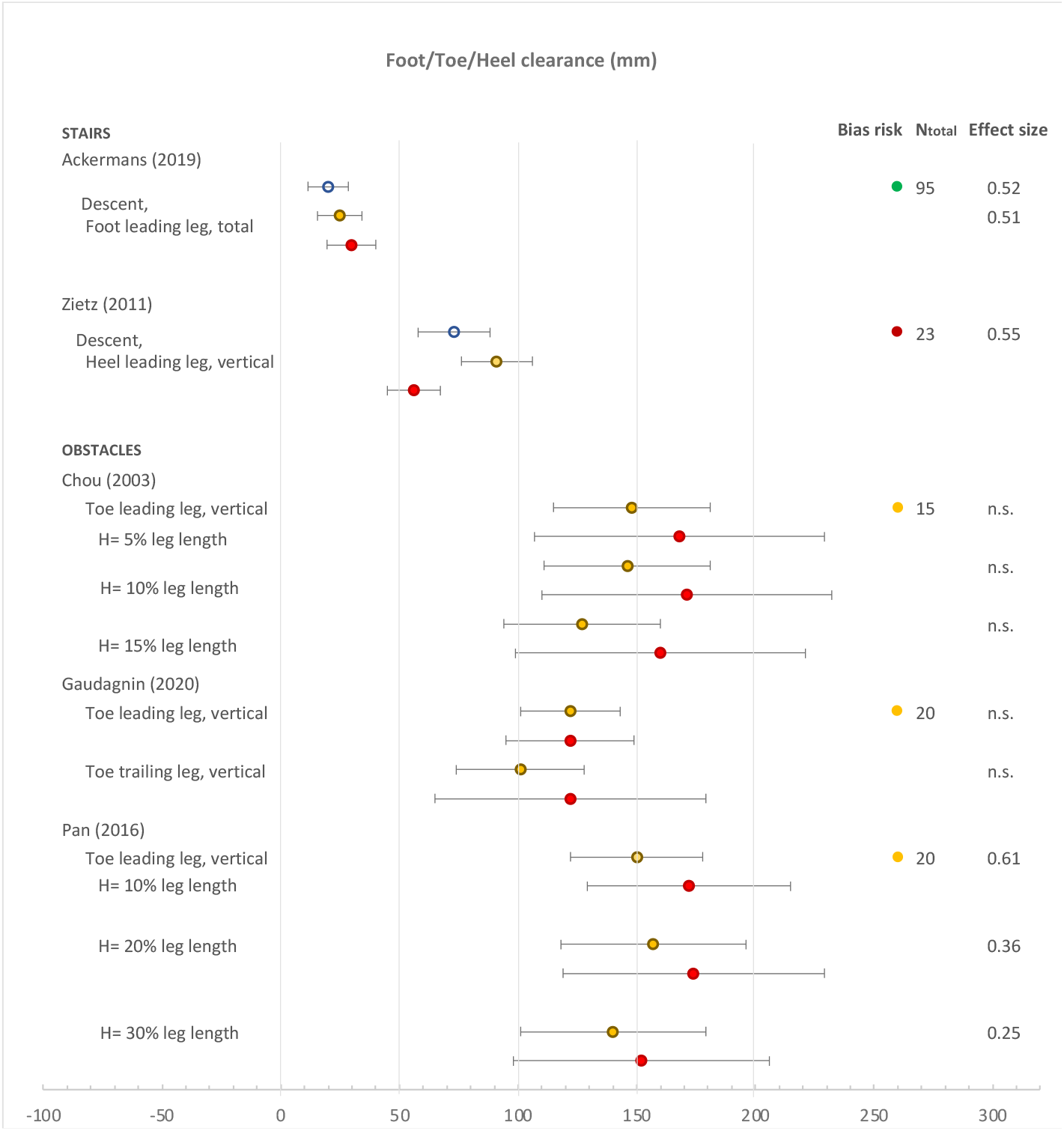
Forest plot of heel/foot/toe clearance. Circles: open blue– younger adults, yellow closed– older adults with low fall risk or non-fallers, red closed – older adults with high fall risk or fallers. Ntot is the total number of participants. n.s. is non-significant finding

Walking speed on stairs was not found to be significantly different between older adults at higher and lower risk when fall risk was assessed prospectively (Ackermans et al., 2021) or retrospectively (Oh-Park et al., 2011; Ackermans et al., 2019). In contrast, older adults at higher risk were found to be slower on stairs when risk was evaluated based on functional decline (Oh-Park et al., 2012) or when comparing a group of older (77.3 ± 7.8 years) hospitalised patients to a group of healthy participants (70.3 ± 5.3 years) (Brodowski et al., 2021). The variance in walking speed was also found to be not significantly different between higher and lower risk older adults (Ackermans et al., 2019, 2021).

#### 3.2.3 Foot clearance

Foot clearance was assessed in 3 risk studies with obstacles and 3 risk studies with stairs, and was found to be not significantly different between older adults at higher and lower risk when fall risk was assessed prospectively (Ackermans et al., 2021), retrospectively (Ackermans et al., 2019; Guadagnin et al., 2020) or based on unsteadiness complaints (Chou et al., 2003). In contrast, subjects with lower physical level have an increased foot clearance on obstacles and reduced symmetry in foot clearance (Pan et al., 2016). On stairs, older subjects (79.3± 6.4 years) with lower physical and mental level have a reduced foot clearance compared to old subjects (72.1 ± 3.8 years) with higher physical and mental level (Zietz et al., 2011). However, variance in foot clearance was not significantly correlated with fall risk assessed either based on retrospective fall history (Ackermans et al., 2019) or physical and mental level (Zietz et al., 2011).

#### 3.2.4 Step length

Step length was assessed in 1 risk study with stairs, 2 with perturbations and 4 with obstacles. It was not found to be significantly different between older adults at higher and lower risk, evaluated based on retrospective fall history (Pieruccini-Faria and Montero-Odasso, 2019; Guadagnin et al., 2020; Gerards et al., 2021), physical and/or mental level (Zietz et al., 2011; Pan et al., 2016), balance performance (Pijnappels et al., 2005) or unsteadiness complaints (Chou et al., 2003) In perturbed walking, one study reported that in trips leading to a fall, the foot was placed backwards of the pelvis during the recovery step, whereas it was placed forwards for trials in which the subjects recovered balance (Pijnappels et al., 2005). The study did not report whether this difference in foot placement was significant. When crossing an obstacle, **step length variability** was found to be larger in older adults with a retrospective fall history (Pieruccini-Faria and Montero-Odasso, 2019), whereas it was not significantly different between groups during unobstructed walking.

#### 3.2.5 Step duration

Stance, swing, initiation, reaction, recovery, or compensatory duration were assessed in 1 risk study with stairs, 2 with perturbations and 3 with obstacles. During stair ascent (but not descent), the support phase was significantly longer for older adults with a lower physical level (Carli et al., 2014). When walking with perturbations, the step duration (Gerards et al., 2021), stance phase and double support durations (Pijnappels et al., 2005) were not significantly different between older adults at higher and lower risk, evaluated based either on retrospective fall history (Gerards et al., 2021) or the subject’s ability to recover their balance after the perturbation (Pijnappels et al., 2005). When crossing an obstacle, step duration increased compared to unobstructed walking, but there was no difference between subjects at higher and lower risk based on retrospective fall history (defined as at least one injurious fall or at least two non-injurious falls) (Pieruccini-Faria and Montero-Odasso, 2019) or unsteadiness complaints (Chou 2003). Subjects with a more severe fall history had a significantly **higher step time variability** when walking over an obstacle, but not during unobstructed walking (Pieruccini-Faria and Montero-Odasso, 2019). When crossing an obstacle, swing time was not significantly different between older adults with higher or lower physical level (Pan et al., 2016). Furthermore, the average stride time and stride time variability when crossing an obstacle course (standing up from chair, walking along narrow path, walking on an uneven surface, crossing 3 obstacles, then either climbing stairs or ending the task) was not significantly different between older adults with and without a prospective fall history (Hansson et al., 2021).

#### 3.2.6 Step width

Step width was assessed in 1 risk study with stairs, 1 with perturbations and 2 with obstacles. When crossing an obstacle, a smaller step width for all obstacle heights was reported for older adults at lower physical level (Pan et al., 2016). In contrast, there was no significant difference in step width between older patients with unsteadiness complaints and healthy controls (Chou 2003). When walking with perturbations, there was no significant difference between older adults with and without a retrospective fall history (Gerards et al., 2021). When climbing stairs, there were no significant differences in step width or step width variability between older adults with a higher and lower physical level (Zietz et al., 2011).

#### 3.2.7 Margin of stability

Margin of stability was assessed in 3 risk studies with stairs and 1 with perturbations, and was found to be not significantly correlated to prospective (Ackermans et al., 2021) or retrospective (Ackermans et al., 2019; Gerards et al., 2021) fall history, or physical and mental level (Zietz et al., 2011).

#### 3.2.8 Outcomes assessed in less than 3 risk studies

When walking on stairs, there were no significant differences in either the percent of the foot surface in contact with the stairs, its variance, or the required coefficient of friction, between older adults at higher and lower risk, evaluated based either on prospective (Ackermans et al., 2021) or retrospective fall history (Ackermans et al., 2019). A cluster analysis combining multiple parameters was able to identify different stair negotiation strategies. However, these strategies could not predict the risk of falling on stairs. For example, older adults with a more conservative strategy for stair descent (i.e. increased foot clearance) have a similar hazard risk to those who adopt a riskier strategy (i.e. reduced foot clearance).

When walking on stairs, subjects with a lower physical level had a significantly lower peak vertical ground reaction force, lower vertical loading and unloading rate, and higher vertical impulse (Carli et al., 2014). Center of mass acceleration and variance were not significantly different between older adults with a higher or lower physical and mental level (Zietz et al., 2011).

Older subjects who fell when they are tripped during walking had **higher angular momentum** compared to those who recovered their balance (Pijnappels et al., 2005). There was however no significant difference in the ankle, knee or hip moments or in hip motion.

When crossing an obstacle, foot placement relative to the obstacle was not significantly different between older adults at higher and lower risk, evaluated based either on retrospective fall history (Guadagnin et al., 2020) or physical level (Pan et al., 2016). Moreover, the medial-lateral foot excursion did not differ between older patients with unsteadiness complaints and healthy controls (Chou 2003). When initiating walking over an obstacle, the **duration of the anticipatory postural adjustments was longer** in older adults with a retrospective fall history (Uemura et al., 2011), but not in unobstructed walking. Lastly, the medial-lateral CoM range of motion and peak velocity were significantly higher in older adults suffering from unsteadiness complaints compared to healthy older adults (Chou 2003). When navigating an obstacle course, **gait flexibility** was reduced in older adults with a prospective fall history (Hansson et al., 2021). In this study, gait flexibility was defined as the difference in the stepping signal from an IMU above the knee between unobstructed walking and navigating the obstacle course.

## 4 Discussion

While many studies have investigated fall risk during steady walking, this task is not comparable to the majority situations in which older people fall in daily life (Tinetti et al., 1988; Speechley and Tinetti, 1991; Luukinen et al., 2000; Mackenzie et al., 2002; Sartini et al., 2010). This is the first systematic review of the performance of younger and older adults with higher and lower fall risk during challenging walking tasks: stair climbing, perturbed walking and obstacle crossing. We identified several motion analysis performance parameters assessed during challenging walking tasks that may be possible candidates to predict the risk of falling in the older population.

### 4.1 Identifying prognostic factors requires measuring fall history

#### 4.1.1 Ageing factors do not predict fall risk

Most of the articles identified by our search strategy (62 out of 78) simply compared healthy younger to healthy older adults. Our results show that the observed differences between younger and older adults are not necessarily good prognostic factors for fall risk in the older population. For example, walking speed is significantly smaller in older than younger adults, but it is not significantly correlated with either prospective (Ackermans et al., 2021) or retrospective (Oh-Park et al., 2011; Ackermans et al., 2019; Pieruccini-Faria and Montero-Odasso, 2019; Guadagnin et al., 2020) fall history. This is consistent with the findings from perturbed stance. There are robust differences in the responses of younger versus older subjects to stance perturbations (Woollacott et al., 1986; Inglin and Woollacott, 1988; Nardone et al., 1995; Allum et al., 2004; Tokuno et al., 2010). However, the response to stance perturbations typically do not differ between older adults with and without a prospective (Baloh et al., 1998; Hill et al., 1999; Kario et al., 2001) or retrospective (Studenski et al., 1991) fall history. Therefore, studies comparing older to younger adults cannot be used to explore relationships between fall risk and motion analysis parameters.

#### 4.1.2 Risk factors in ageing

In the 16 remaining studies which compared two groups of older adults, only 2 studies classified the groups according to prospective fall history, and 6 studies according to retrospective fall history. The remaining 8 studies used a variety of methods to distinguish between higher and lower risk older adults. Importantly, the findings from these latter studies are not always corroborated by the studies which classified older adults according to fall history. For example, walking speed on stairs is not significantly correlated with fall history (Oh-Park et al., 2011; Ackermans et al., 2019, 2021), but it is reduced in subjects who then undergo functional decline (Oh-Park et al., 2012) and in hospitalised patients relative to healthy subjects (Brodowski et al., 2021). Candidate prognostic factors identified from cross-sectional studies which do not measure fall history must therefore be interpreted with caution.

### 4.2 Candidate prognostic factors in challenging locomotion tasks

#### 4.2.1 Factors correlated with prospective fall history

When navigating an obstacle course, **gait flexibility** (the change in stepping pattern relative to unobstructed walking) may be a good prognostic factor for fall risk (Hansson et al., 2021). When walking up and down stairs, no single motion analysis parameter is able to predict subsequent hazard events (including falls) on stairs (Ackermans et al., 2021). However, a **cluster analysis** using several parameters may be useful to identify different stair negotiation strategies (Ackermans et al., 2021). This may be useful to identify how older subjects alter their stair negotiation strategy, either to compensate other deficits, or in response to fear of falling (Anders et al., 2007; Donoghue et al., 2013).

#### 4.2.2 Factors correlated with retrospective fall history

In obstacle crossing, older adults with a retrospective fall history demonstrated a **larger step length variability and step time variability** (Pieruccini-Faria and Montero-Odasso, 2019). When initiating walking over an obstacle, **anticipatory postural adjustments are prolonged** in older adults with a retrospective fall history (Uemura et al., 2011). Prolonged reaction times have also been found to be correlated to prospective fall history when subjects are asked to perform a single step in response to a cue (Melzer et al., 2010; Pijnappels et al., 2010). Prolonged stepping times may however be specific to tasks in which the step is self-initiated in response to an external cue. Indeed, when the step is a response to an external perturbation, stepping initiation is earlier in subjects with a prospective fall history (Mille et al., 2013).

#### 4.2.3 Causes of falling

Most studies focussed on spatiotemporal outcome parameters. Only a limited number of studies tried to relate these parameters and fall risk to other underlying mechanisms such as postural adjustments or angular moment. This may however be a promising avenue for future research. Indeed, as mentioned above, postural adjustments have been related to retrospective fall history (Uemura et al., 2011). Moreover, in one study design, a perturbation was used which caused participants to trip, and fall in a portion of the trials (Pijnappels et al., 2005). This allowed the authors to identify that in trips leading to a fall, the angular momentum was reduced. While spatiotemporal parameters are relatively easy to assess, fall prevention requires a better understanding of the mechanisms underlying poor performance in challenging walking tasks. Measuring the mechanisms which underly falls (such as angular momentum) requires assessing external forces and full-body kinematics with advanced measurement technologies such as load or pressure plates and 3-dimensional movement capture systems.

### 4.3 Measurement technology and setting

All the included studies were performed in a laboratory setting, mostly using traditional motion capture. However, fall risk may be better identified by measuring subjects in their natural environment. Indeed, factors such as ambient lighting, physical or mental fatigue, and stair, obstacle or perturbation type can play a role in increasing fall risk (Startzell et al., 2000; Jacobs, 2016). Inertial Measurement Units (IMU’s) enable ambulant measurements and could be used to monitor stair climbing or obstacle crossing. An IMU sensor on the sacrum (Bolink et al., 2016; Jacobs, 2016; Bartlett and Goldfarb, 2018) or thigh (Hansson et al., 2021) could be used to measure relevant motion parameters, such as gait flexibility, step time or step length variability. Changes in gait initiation can also be detected in ambulatory settings using IMUs (Mancini et al., 2016; King et al., 2017). Such parameters measured during challenging ambulatory tasks could then be incorporated into current fall risk prediction models to improve accuracy, sensitivity, and specificity of fall predictions (Montesinos et al., 2018).

### 4.4 Recommendations

#### 4.4.1 Study design

To determine prognostic factors for falling, fall history must be measured. If possible, this should be done in a longitudinal design with a long-term recording of fall history. When this is not feasible, fall history may alternatively be measured retrospectively. To avoid bias in the results, the subjects must be well described in terms of mental and physical fitness level, and must represent the diversity within the older population living in the community in terms of fall risk, gender, and fitness level. Furthermore, group sample sizes need to be sufficiently large to ensure statistical power.

#### 4.4.2 Motion parameters

The findings suggest that fall risk can be better discriminated from changes or variability in outcome parameters rather than the mean of a given parameter. For example, whereas step time and step length are not correlated with retrospective fall history, their variability is (Pieruccini-Faria and Montero-Odasso, 2019). Similarly, gait flexibility (the change in stepping pattern between unobstructed walking and navigating an obstacle course) is prognostic of falling (Hansson et al., 2021). Moreover, parameters related to the cause of falls such as postural adjustments (Uemura et al., 2011) or angular momentum (Pijnappels et al., 2005) may be relevant to identify fall risk. Also, combinations of parameters may be used to identify the strategies employed by older adults when faced with challenging walking tasks (Ackermans et al., 2019, 2021). Finally, to be able to compare studies in a meta-analysis, in the future motion analysis parameters should be assessed in a uniform way.

### 4.5 Limitations

This review was restricted to ‘healthy’ older adults, i.e., older adults with normal ageing degeneration impairments and no moderate to severe diseases significantly impairing locomotion. Therefore, our results do not apply to older adults suffering from diseases impairing their locomotion such as severe osteoarthritis, stroke or Parkinson disease. Moreover, the review focussed on three challenging walking tasks, which were related to biomechanics risk factors for falling. Other potentially relevant tasks (such as running or dual tasks) were not considered. Due to the task specificity of balance, the findings may not generalise to other tasks.

Finally, although 78 articles were included in this study, fall history was measured in only 8 of these. Moreover, the large variation in assessed outcome parameters resulted in mostly a limited number of studies reporting any given parameter. Therefore, our motion parameter recommendations are based on a very limited number of studies.

## 5 Conclusion

We investigated the relationship between fall risk among the older population and their performance during challenging walking tasks (stair climbing, perturbations and obstacle crossing). The results from the 78 included studies indicated that findings from studies comparing young to older adults cannot be used as prognostic factors for fall risk. Even when comparing two older adult populations, it is necessary to measure fall history so as to identify fall risk prognostic factors. We identified candidate motion analysis factors for fall risk prediction, which could also be assessed ambulatory in a more natural environment. Finally, we provided recommendations for the study design and motion parameters to be assessed in future fall risk assessment studies.

## Supporting information

Appendix A - Search strings

Appendix B - Bias definitions

Appendix C - Risk of bias

Appendix D - Ageing studies

Appendix E - Ageing references

Appendix F - Ageing outcomes

## Data Availability

The study is a systematic review reporting results from only previously published scientific papers.

