## Appendix A - Search strings for "Systematic review of candidate prognostic factors for falling in older adults identified from motion analysis of challenging walking tasks"

### Appendix A: Database search strings

The search engines from Pubmed, IEEExplore and Scopus were used to search for relevant articles. The search string was developed in Pubmed based on our defined participants, context, and outcome measures and then adapted to the other two search engines.

#### 1. Pubmed

**Search string**

("fall" OR "fall risk") AND ("obstacle" OR "stair" OR "perturbation") AND ("age" OR "older" OR "elderly") NOT (diabetes[Title/Abstract]) NOT ("rheumatoid arthritis"[Title/Abstract] OR "osteoarthritis"[Title/Abstract]) NOT (Parkinson[Title/Abstract]) NOT (stroke[Title/Abstract])

Returns 385

#### 2. IEEE

**Search string**

(("All Metadata":"fall" OR "All Metadata":"fall risk") AND ("All Metadata":"obstacle" OR "All Metadata":"stair" OR "All Metadata":"perturbation") AND ("All Metadata":"age" OR "All Metadata":"older" OR "All Metadata":"elderly") NOT ("Document Title":"age" OR "Document Title":"rheumatoid arthritis" OR "Document Title":"osteoarthritis"OR "Document Title":"Parkinson" OR "Document Title":"stroke") )

Returns 49

#### 3. Scopus

**Search string**

( "fall" OR "fall risk" ) AND ( "obstacle" OR "stair" OR "perturbation" ) AND ( "age" OR "older" OR "elderly" ) AND NOT TITLE-ABS ( "diabetes" ) AND NOT TITLE-ABS ( "rheumatoid arthritis" or "osteoarthritis") AND NOT TITLE-ABS ( "Parkinson" ) AND NOT TITLE-ABS ( "stroke" )

Returns 23,050

To limit the number of articles, several modifications have been made to the search string. The different modification steps are given in paragraph 3.1 and the final Scopus search string in paragraph 3.2.

##### 3.1. Modification of the search string for Scopus search

AND ( "obstacle" OR "stair" OR "perturbation" )

Replaced with AND TITLE-ABS ( "obstacle" OR "stair" OR "perturbation" )

Returns: 6,453

AND ("age" OR "older" OR "elderly" )

Replaced with AND ("aging" OR "ageing" OR "older" OR "elderly" )

Returns: 4,854

#### 3.1.1. Domain restrictions

**Relevant domains**:

***Limit to:*** *engineering, health professions, medicine, neuroscience, nursing*

AND ( LIMIT-TO ( SUBJAREA , "MEDI" ) OR LIMIT-TO ( SUBJAREA , "ENGI" ) OR LIMIT-TO ( SUBJAREA , "HEAL" ) OR LIMIT-TO ( SUBJAREA , "NEUR" ) OR LIMIT-TO ( SUBJAREA , "NURS" ) )

Returns: 3,738

***Exclude****: social sciences; physics and astronomy; chemical engineering; materials science; environmental science; immunobiology and microbiology; chemistry; agricultural and biological sciences; pharmacology, toxicology and pharmaceutics; business, management and accounting; arts and humanities; decision sciences; energe; earth and planetary sciences; economics, enconometrics and finance; veterinary; dentistry*

AND ( EXCLUDE ( SUBJAREA,"SOCI" ) OR EXCLUDE ( SUBJAREA,"PHYS" ) OR EXCLUDE ( SUBJAREA,"CENG" ) OR EXCLUDE ( SUBJAREA,"MATE" ) OR EXCLUDE ( SUBJAREA,"ENVI" ) OR EXCLUDE ( SUBJAREA,"IMMU" ) OR EXCLUDE ( SUBJAREA,"CHEM" ) OR EXCLUDE ( SUBJAREA,"AGRI" ) OR EXCLUDE ( SUBJAREA,"PHAR" ) OR EXCLUDE ( SUBJAREA,"BUSI" ) OR EXCLUDE ( SUBJAREA,"ARTS" ) OR EXCLUDE ( SUBJAREA,"DECI" ) OR EXCLUDE ( SUBJAREA,"ENER" ) OR EXCLUDE ( SUBJAREA,"EART" ) OR EXCLUDE ( SUBJAREA,"ECON" ) OR EXCLUDE ( SUBJAREA,"VETE" ) OR EXCLUDE ( SUBJAREA,"DENT" ) )

Returns: 3,412

#### 3.1.2. Language

***Limit to:*** *English*

AND ( LIMIT-TO ( LANGUAGE , "English" ) )

Returns: 3,064

***Exclude****: Portuguese, French, Spanish, Polosh, German, Italian, Japanese, Persian, Turkish*

AND ( EXCLUDE ( LANGUAGE , "Portuguese" ) OR EXCLUDE ( LANGUAGE , "French" ) OR EXCLUDE ( LANGUAGE , "Spanish" ) OR EXCLUDE ( LANGUAGE , "Polish" ) OR EXCLUDE ( LANGUAGE , "German" ) OR EXCLUDE ( LANGUAGE , "Italian" ) OR EXCLUDE ( LANGUAGE , "Japanese" ) OR EXCLUDE ( LANGUAGE , "Persian" ) OR EXCLUDE ( LANGUAGE , "Turkish" ) )

Returns: 3,261

##### 3.1.3. Article type

***Exclude:*** *reviews, books, book series*

AND ( EXCLUDE ( DOCTYPE , "re" ) ) AND NOT TITLE-ABS ("systematic review") AND ( EXCLUDE ( EXACTKEYWORD , " Review" ) AND ( EXCLUDE ( SRCTYPE , "b" ) OR EXCLUDE ( SRCTYPE , "k" ) )

Returns: 2,786

##### 3.1.4. Study design

***Exclude:*** *randomized control trials, clinical trials, interventions, training, treatment*

AND NOT TITLE-ABS ("Randomized Controlled Trial ") AND NOT TITLE-ABS ("Clinical Trial ") AND NOT TITLE ( “intervention” OR “training”) AND EXCLUDE ( EXACTKEYWORD , "Treatment Outcome" ) AND EXCLUDE ( EXACTKEYWORD , "Controlled Clinical Trial" ) AND EXCLUDE ( EXACTKEYWORD , "Randomized Controlled Trial" )

Returns: 2,482

##### 3.1.5. Study population

***Exclude diseases and injuries***: *Alzheimer, Dementia, depression, injury, orthotics, fracture, knee osteoarthritis, sarcopenia*

AND NOT TITLE-ABS ( "Alzheimer" ) AND NOT TITLE-ABS ( "brain injury" ) AND NOT TITLE-ABS ( "Dementia " ) AND NOT TITLE-ABS ( "depression " ) AND NOT TITLE-ABS ( "orthotics" ) AND NOT TITLE ( "injury" ) AND NOT TITLE ( "fracture" ) AND EXCLUDE ( EXACTKEYWORD , "Knee osteoarthritis" ) AND EXCLUDE ( EXACTKEYWORD , "Knee Osteoarthritis" ) AND EXCLUDE ( EXACTKEYWORD , "Sarcopenia" )

Returns: 2,132

***Exclude study populations:*** *non-human animals, pregnancy, children, amputees, obesity*

AND EXCLUDE ( EXACTKEYWORD , "Nonhuman" ) AND NOT TITLE-ABS ( "pregnancy " OR “pregnant” ) AND NOT TITLE-ABS ( "child " OR “children” OR “infant” OR “adolescent”) AND EXCLUDE ( EXACTKEYWORD , "Adolescent" ) AND NOT TITLE-ABS ( "amputation " OR “amputee” ) AND EXCLUDE ( EXACTKEYWORD , "Obesity" )

Returns: 2,027

##### 3.1.6. Task

***Exclude***: *dual task, virtual reality*

AND NOT TITLE-ABS ( "dual task" ) AND ( EXCLUDE ( EXACTKEYWORD , "Virtual Reality" )

Returns: 1,835

#### 3.2. Final search string for Scopus

( "fall" OR "fall risk" ) AND TITLE-ABS ( "obstacle" OR "stair" OR "perturbation" ) AND ("aging" OR "ageing" OR "older" OR "elderly" ) AND NOT TITLE-ABS ( "diabetes" ) AND NOT TITLE-ABS ( "rheumatoid arthritis" or "osteoarthritis") AND NOT TITLE-ABS ( "Parkinson" ) AND NOT TITLE-ABS ( "stroke" ) AND NOT TITLE-ABS ("systematic review") AND NOT TITLE-ABS ("Randomized Controlled Trial ") AND NOT TITLE-ABS ("Clinical Trial ") AND NOT TITLE ( "intervention" OR "training") AND NOT TITLE-ABS ( "Alzheimer" ) AND NOT TITLE-ABS ( "brain injury" ) AND NOT TITLE-ABS ( "Dementia " ) AND NOT TITLE-ABS ( "depression " ) AND NOT TITLE-ABS ( "orthotics" ) AND NOT TITLE ( "injury" ) AND NOT TITLE ( "fracture" ) AND NOT TITLE-ABS ( "dual task" ) AND ( LIMIT-TO ( SUBJAREA,"MEDI" ) OR LIMIT-TO ( SUBJAREA,"ENGI" ) OR LIMIT-TO ( SUBJAREA,"HEAL" ) OR LIMIT-TO ( SUBJAREA,"NEUR" ) OR LIMIT-TO ( SUBJAREA,"NURS" ) OR EXCLUDE ( SUBJAREA,"SOCI" ) OR EXCLUDE ( SUBJAREA,"PHYS" ) OR EXCLUDE ( SUBJAREA,"CENG" ) OR EXCLUDE ( SUBJAREA,"MATE" ) OR EXCLUDE ( SUBJAREA,"ENVI" ) OR EXCLUDE ( SUBJAREA,"IMMU" ) OR EXCLUDE ( SUBJAREA,"CHEM" ) OR EXCLUDE ( SUBJAREA,"AGRI" ) OR EXCLUDE ( SUBJAREA,"PHAR" ) OR EXCLUDE ( SUBJAREA,"BUSI" ) OR EXCLUDE ( SUBJAREA,"ARTS" ) OR EXCLUDE ( SUBJAREA,"DECI" ) OR EXCLUDE ( SUBJAREA,"ENER" ) OR EXCLUDE ( SUBJAREA,"EART" ) OR EXCLUDE ( SUBJAREA,"ECON" ) OR EXCLUDE ( SUBJAREA,"VETE" ) OR EXCLUDE ( SUBJAREA,"DENT" ) ) AND ( EXCLUDE ( DOCTYPE,"re" ) ) AND ( LIMIT-TO ( LANGUAGE,"English" ) OR EXCLUDE ( LANGUAGE,"Portuguese" ) OR EXCLUDE ( LANGUAGE,"French" ) OR EXCLUDE ( LANGUAGE,"Spanish" ) OR EXCLUDE ( LANGUAGE,"Polish" ) OR EXCLUDE ( LANGUAGE,"German" ) OR EXCLUDE ( LANGUAGE,"Italian" ) OR EXCLUDE ( LANGUAGE,"Japanese" ) OR EXCLUDE ( LANGUAGE,"Persian" ) OR EXCLUDE ( LANGUAGE,"Turkish" ) ) AND ( EXCLUDE ( EXACTKEYWORD,"Review" ) OR EXCLUDE ( EXACTKEYWORD,"b OR EXCLUDE SRCTYPE" ) OR EXCLUDE ( EXACTKEYWORD,"Treatment Outcome" ) OR EXCLUDE ( EXACTKEYWORD,"Controlled Clinical Trial" ) OR EXCLUDE ( EXACTKEYWORD,"Randomized Controlled Trial" ) OR EXCLUDE ( EXACTKEYWORD,"Knee osteoarthritis" ) OR EXCLUDE ( EXACTKEYWORD,"Knee Osteoarthritis" ) OR EXCLUDE ( EXACTKEYWORD,"Sarcopenia" ) OR EXCLUDE ( EXACTKEYWORD,"Nonhuman" ) OR EXCLUDE ( EXACTKEYWORD,"f NOT TITLE-ABSchild ORchildren ORinfant ORadolescent" ) OR EXCLUDE ( EXACTKEYWORD,"Adolescent" ) OR EXCLUDE ( EXACTKEYWORD,"f EXCLUDE EXACTKEYWORD" ) OR EXCLUDE ( EXACTKEYWORD,"Virtual Reality" ) )
