## Appendix B - Bias definitions for "Systematic review of candidate prognostic factors for falling in older adults identified from motion analysis of challenging walking tasks"

Table B.1: Overview of the bias domains with corresponding signalling questions, sub-questions and context-specific examples.

| **BIAS domain &**<br><br>**main signalling question** | **Sub-questions** | **Context examples** |
|---|---|---|
| **1. Confounding factors** | *i.e,. if another factor influences both the fall risk assessment and the kinematic measurement* | |
| 1.1 Is there potential for confounding of the effect of group allocation? | 1.i Are confounding factors measured?<br><br>1.ii Were the confounding factors measured with valid and reliable tools? | Unintentional differences in groups for age, gender, frailty, mental and physical fitness, or history of falling. |
| | 1.iii Were the confounding factors similar in all groups?<br><br>1.iv. If not, was an appropriate analysis method used to control for confounding factors? | E.g., confounding when the older participant groups are not age-matched? |
| **2. Participant selection** | *i.e., the study sample adequately represents the population of interest* | |
| 2.1 Was selection of participants into the study based on participant characteristics of the target population?<br><br>So, is there adequate participation into the study by eligible persons? | 2.i Are the participants adequately described? Baseline study sample, recruitment, inclusion and exclusion criteria<br><br>2.ii Do the participant characteristics match those of the target population?<br><br>2.iii Are there unjustified exclusions? | i. Characteristics of target population:<br><br>- Age: “older” group should have a mean age of at least 65<br>- Gender: half females, half males<br>- Health status: no systemic diseases<br>- Living in the community (not in care homes, as subjects living in care homes may have less exposure to challenged walking tasks)<br>iii. Unjustified exclusions are e.g.:<br><br>-Low physical and mental fitness<br>-Previous history of falls |

| | | |
|---|---|---|
| 2.2 In case of prospective studies: Does the time between experiment and fall event coincide for most participants? | 2.iv Is the follow-up time the same for all participants? | |
| **3. Group allocation based on fall risk assessment** | *i.e., the participants are correctly allocated to high and low fall risk groups* | |
| 3.1 Were fall-risk groups clearly defined? | 3.i Is the method applied in the same way to all participants?<br><br>3.ii Are the assessment methods valid and reliable fall-risk assessment methods? | ii. Recall of fall occurrence; If fall risk assessment is based on the subjects reporting falls (either prospectively or retrospectively): were they asked to recall falls occurring more than 6 months beforehand? Note that subjects with low mental status may recall falling less accurately. |
| 3.2 Was the information used to define groups recorded before the experiment? | 3.ii If fall risk is assessed after the experiment, could it be influenced by the performance during the experiment? | |
| 4**. Intended experiment** | *i.e., the experiment was performed in a similar manner for all participants* | |
| 4.1 Were their deviations in intended experiment between the groups? | 4.i Could the experiment be influenced by assessor knowledge of the fall risk assessment?<br><br>4.ii Were these deviations from intended experiment unbalanced between groups and likely to have affected the outcome? | i. Were assessors blinded to the population group?<br><br>- Age: blinding is not possible<br>- Balance measurement: was this done before or after the kinematic experiment? If it was done before, where the assessors aware of the outcome?<br><br>ii. Effect of instructions: Differences in dis- or encouragements during experiment or in explanation of task details between groups.<br><br>E.g., pointing out harness usage or handrail |

| | | |
|---|---|---|
| 4.2 Was the experiment implemented successfully for most participants? | 4.ii Were all participants offered the same experiment? | ii. Were the trials presented in random order? |
| 4.3 Were study participants able to follow and adhere to the assigned experiment? | 4.iii Was an appropriate analysis used to estimate the effect of implementing and adhering to the experiment? | iii. Where the subjects allowed to rest to prevent fatigue?<br><br>iv. Was there a difference across groups in the number of participants unable to perform the task (for example: taking the stairs without using the handrails)? |
| **5. Motion analysis outcome parameters** | *i.e., the outcome parameters are measured in similar way for all participants* | |
| 5.1 Could the outcome parameters be influenced by assessor knowledge of the fall risk assessment (group allocation)? | 5.i If so, were assessors aware of the fall risk assessment of the study participants?<br><br>5.ii Were outcome parameters subjectively assessed? | ii. Were assessors blinded to population group?<br><br>- Age: blinding is not possible<br>- Fall history<br>- Physical or mental fitness |
| 5.2 Were the methods of parameter assessment comparable across intervention groups? | 5.iii Clear definition of outcome parameters provided?<br><br>5.iii Methods are adequately valid and reliable, method and setting similar for all participants? | Did all participants receive the same outcome parameter assessment methods and thresholds, at same time point, with same definition, and same measurements? |
| 5.3 Were any systematic errors in outcome parameters related to fall risk group? | | Systematic measurement errors across groups, e.g., CoM assessment by means of ‘only’ sacrum markers |
| **6. Missing data** | *i.e., the study data available represent the study sample* | |

| | | |
|---|---|---|
| Are the outcome parameters available for (nearly) all participants? i.e., …<br><br>6.1 Is the fall risk assessment available for all participants? | If potential bias….<br><br>6.i Reasons for missing data and adequate description of corresponding missing participants?<br><br>6.ii. Are the proportion of participants and reasons for missing data provided? Are they similar across fall risk groups? | i. Is there the same proportion of missing data across groups?<br><br>ii. If not:<br><br>- If the data is missing due to technical failure, this would not be expected to have a large influence on the results.<br>- If the data is excluded based on task performance, this can cause bias (e.g., data excluded from analysis if participants grabbed the handrail during the task or because the participant was not able to perform task) |
| 6.2 Were participants excluded due to missing data on other variables needed for the analysis? | 6.iii Is there evidence that results were robust to the presence of missing data? | |
| **7. Result reporting** | *i.e., analysis is appropriate and primary outcomes reported* | |
| Is the reported effect estimate likely to be selected, on the basis of the results, from…<br><br>7.1. … multiple outcome parameters within one outcome domain? | 7.i Is selected statistical method adequate for the design of the study? | i. Did the analyst perform a statistical analysis?<br><br>- If yes, is the statistical method described?<br><br>- if not, is the reported difference between or within groups significantly different based on reported means and corresponding standard deviations (using G*power)? |
| 7.2 … multiple analyses of the fall risk – outcome parameter relationship? | 7.ii Is there selective reporting? | ii. Did the analyst report all generated estimates or only part of the assessed outcome parameters? |
| 7.3 ... different subgroups? | | |

Table B.2: Definition of low, moderate and high risk for the seven bias domains

| | Low | Moderate | High |
|---|---|---|---|
| 1 | The groups compared are matched for gender, age and status. | There is a moderate difference across groups in either gender, age or status. | There is either a large difference in one factor or a difference in several factors. |
| 2 | The overall study population has a balanced gender ratio (between 40 % and 60 % males) AND there are no inappropriate exclusions. | The gender balance in the overall study population is unbalanced ( < 40 % of either males or females)<br><br>AND/OR<br><br>Subjects are excluded based on status (physical health, mental health, fall history). | A single gender is studied<br><br>AND<br><br>Subjects are excluded based on status (physical health, mental health, fall history). |
| 3 | High and low risk older adults are differentiated based on fall history (either prospective or retrospective) | High and low risk older adults are differentiated based on valid clinical tests or questionnaires. | High and low risk older adults are differentiated based on a criterion which is not correlated with fall risk. |
| 4 | The trial order is either randomised or identical for the different groups.<br><br>AND<br><br>Subjects were allowed to take time to recover from fatigue between trials.<br><br>AND<br><br>All groups were able to perform the task. | The trial order is different for the different groups.<br><br>OR<br><br>Subjects were not able to recover from fatigue between trials.<br><br>OR<br><br>The task was too difficult for one of the groups. | 2 of the 3 issues are present. |

| | | | |
|---|---|---|---|
| 5 | All parameters are well defined and valid. | Parameters are well defined, but there is a bias in parameter outcomes based on the group characteristics. | Parameters are not defined. |
| 6 | There is no missing data.<br><br>OR<br><br>There is a small amount of missing data due to technical failure which would not have influenced the results. | There is a lot of missing data due to technical failure.<br><br>OR<br><br>A small amount of data is selectively removed from the analysis based on task performance itself. | Data is selectively removed from the analysis based on task performance itself.<br><br>AND<br><br>A difference in task performance between groups led to a large amount of data being removed from the analysis for one of the groups. |
| 7 | The significance level of all assessed variables is reported. | The significance level of one or several variables is not reported, but the sample size is sufficient for the reported means and standard deviations (see G*power calculation in Methods). | The significance level of one or several variables is not reported, and the sample size is too low. |
