## Appendix C - Risk of bias for "Systematic review of candidate prognostic factors for falling in older adults identified from motion analysis of challenging walking tasks"

**Appendix C: Risk of bias assessment for each of the studies**

Abbreviations: m: male; f: female; No information on status: no assessment of the participants' physical health, mental health or fall history

**Table C.1** Risk of bias assessment for the studies comparing higher and lower risk older adults

| Bias domain | Total bias | 1 – Confounding factors | 2 – Participant selection | 3 – Group allocation based on fall risk assessment | 4 – Intended experiment | 5 – Motion analysis outcome parameters | 6 – Missing data | 7 – Result reporting |
|---|---|---|---|---|---|---|---|---|
| **Stairs** | | | | | | | | |
| Ackermans (2019) | Low | No information on gender. | No information on gender. | Low<br>Retrospective fall history | Low<br>Subjects were allowed to take breaks to avoid fatigue. | Low | Low | Low |
| Ackermans (2021) | Moderate | No information on gender balance within groups. | Moderate<br>Gender is unbalanced: 28 m, 53 f | Low<br>Prospective fall history | Low | Low | Low | Low |
| Brodowski (2021) | Serious | Serious<br>Participants in the hospitalized group were significantly older than in the healthy group ($p < 0.001$, mean age difference 7 years). | Moderate<br>Gender is unbalanced: 43 m, 70 f<br>3 of the 60 hospitalized subjects were excluded because they could not perform the timed-up-and-go test. | Moderate<br>Health status (hospitalized versus healthy) | Low | Low | Moderate<br>4 of the 60 healthy participants were excluded because they performed the task very rapidly or very slowly.<br>4 of the 56 hospitalized subjects could not perform the 2nd trial. | Moderate<br>Significance level was not reported, but sample size was sufficient for the reported effect size. |
| Carli (2014) | Moderate | No information on gender balance within groups. | Moderate<br>Gender is unbalanced: 4 m, 30 f. | Moderate<br>Physical level | No information on fatigue prevention strategy. | Low | Low | Low |

| | | | | | | | | |
|---|---|---|---|---|---|---|---|---|
| Oh-Park (2012) | Moderate | Low | Moderate<br>Gender is unbalanced: 147 m, 224 f. | Moderate<br>Functional decline over 1 year | Low<br>Subjects were allowed a brief rest. | Low | Low | Moderate<br>Significance level was not reported, but sample size was sufficient for the reported effect size. |
| Oh-Park (2011) | Moderate | No information on gender balance within groups | Moderate<br>Gender is unbalanced: 204 m & 309 f. | Low<br>Retrospective fall history | Low<br>Subjects were allowed a brief rest. | Low | Low | Low |
| Zietz (2011) | Serious | Serious<br>Higher risk participants were significantly older ($p < 0.05$, mean age difference 7 years). | Moderate<br>Gender is unbalanced: 3 m, 19 f | Moderate<br>Physical and mental level | Moderate<br>4 out of the 7 high fall risk older adults (and no other participant) used the handrail.<br>No information on fatigue prevention strategy. | Serious<br>No information on how the center of mass is calculated. | Low | Low |
| **Perturbations** | | | | | | | | |
| Gerards (2021) | Low | Low | Low | Low<br>Retrospective fall history | No information on randomization.<br>No information on fatigue prevention strategy. | Low | Low<br>One subject was excluded because of technical failure | Low |
| Pijnappels (2005) | Moderate | Low | No information on status. | Moderate<br>Physical level | No information on fatigue prevention strategy.<br>No information on randomization. | Low | Moderate<br>Older adults (but no younger adults) sometimes performed “a lowering strategy at mid-swing”, and these trials were | Low |

| | | | | | | | | |
|---|---|---|---|---|---|---|---|---|
| | | | | | | | removed from the analysis. | |
| **Obstacles** | | | | | | | | |
| Brach (2011) | Moderate | Low | Moderate<br>Gender is unbalanced: 20 m, 87 f<br>Exclusion of subjects with low walking speed (36 out of the 120 recruited subjects). | Low<br>Retrospective fall history | No information on randomization.<br>No information on fatigue prevention strategy. | Low | Moderate<br>13 out of 84 subjects did not complete the follow-up. | Low |
| Chou (2003) | Moderate | Moderate<br>Gender balance is different across groups<br>Y: 7 m, 2 f<br>O: 1 m, 5 f | Low | Low | Low | Low | Low | Low |
| Hansso n (2021) | Moderate | Low | Moderate<br>Gender is unbalanced: 10 m, 91 f | Low | Low | Low | Low | |
| Guada gnin (2020) | Moderate | Low | Moderate<br>Exclusion of male subjects. | Low<br>Retrospective fall history | No information on fatigue prevention strategy.<br>No information on randomization. | Low | Low | Low |
| Pan (2016) | Moderate | No information on gender. | No information on gender. | Moderate<br>Physical level | Low<br>Subjects were allowed to rest if needed. | Low | Low | Low |
| Pierucc ini- | Moderate | Low | Moderate<br>Exclusion of male subjects. | Low<br>Retrospective fall history | No information on fatigue prevention strategy. | Low | Low | Low |

| | | | | | | | | |
|---|---|---|---|---|---|---|---|---|
| Faria (2019) | | | | | No information on randomization. | | | |
| Uemura (2011) | Moderate | Low | Moderate<br>Gender is unbalanced: 26 m, 50 f. | Low<br>Retrospective fall history | No information on fatigue prevention strategy. | Low | Low | Low |

**Table C.2** Risk of bias assessment for the studies comparing younger and older adults

| Bias domain | Total bias | 1 – Confounding factors | 2 – Participant selection | 4 – Intended experiment | 5 – Motion analysis outcome parameters | 6 – Missing data | 7 – Result reporting |
|---|---|---|---|---|---|---|---|
| **Stairs** | | | | | | | |
| Begg (2000) | Moderate | Low | Moderate<br>Exclusion of male subjects. | No information on fatigue prevention strategy. | Low | Low | Low |
| Bosse (2012) | Low | Low | No information on status. | No information on randomization.<br>No information on fatigue prevention strategy. | Low | Low | Low |
| Chiu (2015) | Low | Low | No information on status. | No information on randomization.<br>No information on fatigue prevention strategy. | Low | Low | Low |
| Christina (2002) | Moderate | Moderate<br>Gender balance is different across groups<br>Y: 5 m, 7 f<br>O: 8 m, 4 f | Low | No information on fatigue prevention strategy. | Low | Low<br>1 participant slipped and was removed from the analysis. | Low |

| | | | | | | | |
|---|---|---|---|---|---|---|---|
| Crosbie (2003) | Low | Low | Low | Low | Low | Low | Low |
| Dewolf (2021) | Moderate | Moderate<br>Gender balance is different across groups<br>Y: 4 m, 4 f<br>O: 9 m, 1 f | Moderate<br>Gender is unbalanced: 13 m, 5 f | Low | Low | Low | Low |
| Dixona (2018) | Low | No information on gender. | Low | No information on randomization.<br>No information on fatigue prevention strategy. | Low | Low | Low |
| Francksen (2022) | Low | No information on gender. | No information on status or gender. | Low | Low | Low<br>2 out of 27 younger adults and 1 out of 33 older adults were excluded<br>due to technical issues. | Low |
| Francksen (2020) | Low | No information on gender. | No information on status or gender. | No information on randomization.<br>No information on fatigue prevention strategy. | Low | Low | Low |
| Foster (2019) | Low | Low | No information on status. | No information on randomization.<br>No information on fatigue prevention strategy. | Low | Low | Low |
| Hamel (2005) | Moderate | Low | Moderate<br>Exclusion of male subjects. | No information on randomization.<br>No information on fatigue prevention strategy. | Low | Low<br>Only trials of a pre-specified speed were included in the analysis. | Low |

| | | | | | | | |
|---|---|---|---|---|---|---|---|
| Hsue (2014) | Moderate | Low | Moderate<br>Exclusion of subjects with fall history. | Low | Low | Low | Low |
| Hsue (2009) | Moderate | Low | Moderate<br>Exclusion of male subjects. | Low | Low | Low | Low |
| Kim (2009) | Moderate | Low | Moderate<br>Exclusion of subjects with low physical test scores. | No information on randomization.<br>No information on fatigue prevention strategy. | Low | Low | Low |
| Larsen (2008) | Moderate | No information on gender. | Moderate<br>Exclusion of subjects who do not participate in physical activities at least once per week.<br>No information on gender. | No information on randomization.<br>No information on fatigue prevention strategy. | Low | Low<br>1 out of 19 older subjects was excluded because they could not walk up the stairs without the handrails. | Low |
| Mian (2007) | Moderate | Low | Moderate<br>Exclusion of female subjects. | No information on randomization.<br>No information on fatigue prevention strategy. | Low | Low | Low |
| Novak (2011) | Moderate | Low | Moderate<br>Exclusion of subjects with fall history.<br>Gender is unbalanced<br>21 m, 36 f. | No information on fatigue prevention strategy. | Low | Low<br>1 subject per group was excluded due to technical failure. | Low |
| Novak (2016) | Moderate | Low | No information on status. | Low<br>Trials were randomized to prevent fatigue affecting the results. | Moderate<br>Markers placed on the sacrum were used as a proxy for the | Low | Low |

| | | | | | | | |
|---|---|---|---|---|---|---|---|
| | | | | | center of mass. | | |
| Reeves (2007) | Moderate | Moderate<br>Gender balance is different across groups<br>Y: 10 m, 7 f<br>O: 5 m, 10 f | No information on status. | No information on randomization.<br>No information on fatigue prevention strategy. | Low | Low | Low |
| **Perturbations** | | | | | | | |
| Afschrift (2019) | Low | No information on gender. | No information on gender. | Low | Low | Low | Low |
| Bosquée (2021) | Low | No information on gender. | No information on status or gender. | Low | Low | Low | Low |
| Debelle (2021) | Moderate | Moderate<br>Gender balance is different across groups<br>Y: 8 m, 9 f<br>O: 3 m, 14 f | Moderate<br>Gender is unbalanced: 11 m, 23 f<br>No information on status. | Low | Low | Low<br>Trials for which the subjects required more than 15 recovery steps were not analyzed (9 younger adults trials and 9 older adults trials) | Low |
| Jeon (2022a) | Moderate | Moderate<br>Gender balance is different across groups<br>Y: 4 m, 6 f<br>O: 7 m, 3 f | Low | Low | Low | Moderate<br>1 out of 10 young subjects was excluded because they had no arm reaction. | Low |

| | | | | | | | |
|---|---|---|---|---|---|---|---|
| Jeon (2022b) | Moderate | Low | No information on status | No information on randomization.<br>No information on fatigue prevention strategy. | Low | Low | Low |
| Kazanski (2020) | Moderate | Low | Low | Low<br>There was an approximately 2 minute rest between trials. | Low | Moderate<br>2 of the 17 older adults (and none of the younger adults) used the handrail, and were removed from the analysis. | Low |
| Laudani (2021) | Moderate | Low | Moderate<br>Gender is unbalanced: 4 m, 16 f .<br>Exclusion of highly active individuals. | Low | Low | Low | Low |
| Liu (2009) | Moderate | No information on gender. | No information on status or gender. | No information on randomization.<br>No information on fatigue prevention strategy. | Low | Moderate<br>"Participants who successfully recovered with two feet (i.e., stance foot and swing foot) on one force-plate were excluded from the current study due to the difficulty in performing inverse dynamics analyses." | Low |
| Martelli (2017) | Moderate | Low | Moderate<br>Exclusion of subjects with fall history. | Low<br>Subjects were allowed to rest if needed.<br>No information on randomization. | Low | Low<br>2 out of 24 trials (both in elderly subjects) were removed due to technical failure. | Low |

| | | | | | | | |
|---|---|---|---|---|---|---|---|
| McCrum (2016) | Moderate | Low | Moderate<br>Exclusion of male subjects.<br>No information on status | No information on randomization.<br>No information on fatigue prevention strategy. | Low | Low | Low |
| McIntosh (2016) | Moderate | Low | Moderate<br>Gender is unbalanced: 6 m, 15 f | Low | Low | Low | Low |
| Nachmani (2020) | Serious | No information on gender. | No information on gender. | Low<br>Subjects were allowed to rest if needed. | Moderate<br>The center of mass was calculated as the midpoint between both ASIS markers. | Serious<br>13 of the 35 older adults (and none of the younger adults) failed to complete the task and were removed from the analysis. | Low |
| Qiao (2018) | Moderate | Low | Moderate<br>Exclusion of subjects with fall history or BMI > 30. | Low | Low | Low | Low |
| Ren (2022) | Moderate | No information on gender. | Moderate<br>Exclusion of subjects with fall history.<br>No information on gender. | Low | Low | Low | Low |
| Roeles (2018) | Moderate | Moderate<br>Gender balance is different across groups<br>Y: 6 m, 3 f<br>O: 2 m, 7 f | Low | No information on fatigue prevention strategy. | Low | Low | Low |
| Rum (2020) | Moderate | Low | Moderate<br>Gender is unbalanced: 4 m, 16 f | Low | Low | Low | Low |

| | | | | | | | |
|---|---|---|---|---|---|---|---|
| Shulman (2018) | Low | Low | Low | No information on fatigue prevention strategy. | Low | Low | Low |
| Shulman (2019) | Low | Low | No information on status. | No information on fatigue prevention strategy. | Low | Low<br>4 of the 560 trials (all in older adults) were removed from the analysis because participants contacted the floor with the forefoot prior to heel contact. | Low |
| Tropea (2015) | Serious | Moderate<br>Gender balance is different across groups<br>Y: 4 m, 2 f<br>O: 2 m, 4 f | Low<br>Exclusion of participants with low physical health (as assessed by a physical therapist) | No information on fatigue prevention strategy.<br>No information on randomization. | Low | Low | Serious<br>Significance level was not reported, and sample size was too low for the reported effect size. |
| Yoo (2021) | Moderate | Moderate<br>Gender balance is different across groups<br>Y: 7 m, 7 f<br>O: 4 m, 10 f | Moderate<br>Gender is unbalanced: 11 m, 17 f | Low | Low | Low | Low |
| **Obstacles** | | | | | | | |

| | | | | | | | |
|---|---|---|---|---|---|---|---|
| Caetano (2016) | Moderate | Low | Moderate<br>Gender is unbalanced: 25 m, 46 f. | No information on fatigue prevention strategy. | Low | Moderate<br>22% of the older adults (and none of the younger adults) made at least 1 mistake, and trials with a mistake were removed from the analysis. | Low |
| Chen (1991) | Low | Low | Low | No information on fatigue prevention strategy. | Low | Low | Low |
| Chen (1994) | Moderate | Low | Low | No information on fatigue prevention strategy. | Low | Moderate<br>Trials in which participants failed to step over an obstacle were removed from the analysis. There is no information on how often this occurred. | Low |
| Chien (2018) | Moderate | Low | Moderate<br>Exclusion of subjects with fall history | Low<br>Subjects rested one minute between trials. | Low | Low | Low |
| Draganich (2004) | Low | Low | No information on status. | Low | Low | Low | Low |
| Eyal (2020) | Moderate | Low | Moderate<br>Exclusion of subjects with fall history. | Low | Low | Low | Low |
| Hahn (2004) | Low | Low | Low | No information on randomization.<br>No information on fatigue prevention strategy. | Low | Low<br>19 out of 6630 individual data points were removed because they were | Low |

| | | | | | | | |
|---|---|---|---|---|---|---|---|
| | | | | | | "outside reasonable variability" | |
| Huang (2008) | Low | No information on gender. | No information on status or gender. | No information on randomization.<br>No information on fatigue prevention strategy. | Low | Low | Low |
| Kim (2013) | Low | Low | Low | No information on fatigue prevention strategy. | Low | Low | Low |
| Kulkarni (2021) | Moderate | Low | Moderate<br>Gender is unbalanced: 7 m, 24 f<br>No information on status. | No information on randomization.<br>No information on fatigue prevention strategy. | Low | Low | Low |
| LoJacono (2018) | Moderate | No information on gender. | No information on gender. | Low | Low | Low | Low |
| Lowrey (2007) | Low | Low | Low | Low | Low | Low | Low |
| Lu (2006) | Low | No information on gender. | No information on status or gender. | No information on fatigue prevention strategy.<br>No information on randomization. | Low | Low | Low |
| Luo (2022) | Serious | Low | Serious<br>Exclusion of:<br>• subjects with fall history.<br>• female subjects. | Low | Low | Low | Low |
| Maidana (2018) | Low | Low | Low | Low | Low | Low | Low |
| McFadyen (2002) | Moderate | Low | Moderate<br>Exclusion of female subjects. | Low | Low | Low | Low |

| | | | | | | | |
|---|---|---|---|---|---|---|---|
| Mckenzie (2004) | Moderate | Low | Moderate<br>Gender is unbalanced: 12m, 20 f | No information on fatigue prevention strategy. | Low | Moderate<br>6 out of 17 older adults and 8 out of 15 younger adults were not analyzed due to technical failure. | Low |
| Park (2012) | Moderate | Low | Moderate<br>Gender is unbalanced: 6 m, 12 f | Low | Low | Low | Low |
| Tomar (2019) | Serious | Low | Moderate<br>Exclusion of female subjects. | No information on randomization.<br>No information on fatigue prevention strategy. | Low | Low | Serious<br>Significance level was not reported, and sample size was too low for the reported effect size. |
| Uchiyama (2012) | Low | No information on gender. | Low<br>No information on gender. | Low | Low | Low | Low |
| Wang (2010) | Low | Low | Low | No information on fatigue prevention strategy.<br>No information on randomization. | Low | Low | Low |
| Weerdesteyn (2005a) | Moderate | Low | Moderate<br>Gender is unbalanced: 26 m, 98 f<br>Exclusion of subjects without fall history. | No information on fatigue prevention strategy. | Low | Low<br>1% of trials were removed from the analysis because the subject performed an | Low |

| | | | | | | | |
|---|---|---|---|---|---|---|---|
| | | | | | | "intermediate avoidance strategy". | |
| Weerdesteyn (2005b) | Serious | Low | Serious<br>Exclusion of male subjects. Subjects were recruited from elderly sports groups. | Low | Low | Low | Low |
