## Appendix D - Ageing studies for "Systematic review of candidate prognostic factors for falling in older adults identified from motion analysis of challenging walking tasks"

**Appendix D - Description of the ageing studies, ordered by walking task.**

Abbreviations: years old: y; male: m; female: f.

Shading: not reported

Risk of bias: ● Low, ● Moderate, ● Serious

Assessment of participant status:

- Fall history:
  - ✓ if fall history preceding the measurement was assessed
  - × if fall history was assessed and fallers excluded from the study; in this case, we indicate the older adults as being at low fall risk.
- Physical/mental level:
  - ✓ if physical/mental level was tested
  - × if physical/mental level was tested and subjects with low (respectively high) fitness were excluded from the study; in this case, we indicate the older adults as being at low (respectively high) risk.

Medicine usage

- ✓ if participants took medication during the measurement period
- × if participants did not take medication during the measurement period

Measurement devices: if force platforms or infrared cameras were used, but their number was not reported, this is indicated by ✓

Task description: Expected:

- × if unexpected changes in the task occurred from trial to trail (for example: if various obstacles or stairs were used and these were presented in random order, or if perturbations occurred at random times)
- ✓ otherwise

| Article | Risk of bias | Assessment of participant status | Participants | Measurement devices | Task description |
|---|---|---|---|---|---|

| | | Fall history (retrospective) | Physical level | Mental level | Community dwelling | Medicine usage | Younger adults | Older adults | Older adults at higher risk | Older adults at lower risk | Force platform | Infrared cameras system | Task | Expected | Self-selected walking velocity | Randomization or fatigue avoidance |
|---|---|---|---|---|---|---|---|---|---|---|---|---|---|---|---|---|
| **Stairs** | | | | | | | | | | | | | | | | |
| Begg (2000) | | | × Questionnaire of musculoskeletal and visual impairments | | ✓ | | 6 (21.2 ± 2 y) (All females) | 6 (67.6 ± 4.8 y) (All females) | | | | ✓ | Stepping onto raised surface Height: 15 cm Length: 500 cm Width: 100 cm | ✓ | Self-selected | ✓ |
| Bosse (2012) | | | | | ✓ | | 13 (25 ± 2 y) (6 m & 7 f) | 13 (69 ± 4 y) (7 m 6 f) | | | 3 | 13 | Descent 2 steps, each Height: 17 cm Width: 30 cm | ✓ | Self-selected | |
| Chiu (2015) | | | | | | | 20 (25 ± 4.5 y) (10 m & 10 f) | 20 (74.3 ± 5.9 y, 10 m & 10 f) | | | | 12 | Descent + ascent 4 steps, each Height: 17 cm Width: 28 cm | ✓ | Self-selected | |
| Christina (2002) | | | ✓ Visual acuity | | | × | 12 (24 ± 3.3 y) (5 m & 7 f) | 12 (73.3 ± 1.9 y (8 m & 4 f) | | | 2 | | Descent + ascent 7 steps, each Height: 18 cm Width: 28 cm | ✓ | Fixed velocity 0.65 m/s ± 0.04 m/s | ✓ |
| Crosbie (2003) | | | ✓ Melbourne Edge test | | ✓ | | 6 (21.9 y) (3 m & 3 f) | 15 (73.1 y) (7 m & 8 f) | | | | 1 | Stepping onto and off a raised surface | ✓ | Self-selected | ✓ |

| | | | | | | | | | | | | | | | | |
|---|---|---|---|---|---|---|---|---|---|---|---|---|---|---|---|---|
| | | | | | | | | | | | | | Height: 9 cm<br>Length: 35 cm<br>Width: 100 cm | | As fast as possible | |
| Dewolf (2021) | | ✓ | | | | | 8<br>(28.4 ± 5.2 y)<br>(4 m & 4 f) | | | 10<br>(73.5 ± 4.5 y)<br>(9 m & 1 f) | | 9 | Descent + ascent<br>2 steps, each<br>Height: 13 cm<br>Width: 24.5 cm | ✓ | Self-selected | ✓ |
| Dixon (2018) | | | ✓<br>Timed-Up-and-Go | | | | 20<br>(24 ± 3 y) | 19<br>(74.2 ± 6 y) | | | 4 | 12 | Descent + ascent<br>4 steps | ✓ | | |
| Francksen (2022) | | | | | | | 27<br>(24 ± 3 y) | 33<br>(70 ± 4 y) | | | 4 | ✓ | Descent + ascent<br>7 steps, each<br>Height: 20 cm<br>Width: 24 to 26 cm | × | | ✓ |
| Francksen (2020) | | | | | ✓ | | 26<br>(24 ± 3y)<br>No gender information | 33<br>(70 ± 4y)<br>No gender information | | | 4 | 23 | Descent + ascent<br>7 steps, each<br>Height: 20 cm<br>Width: 24 to 26 cm | × | | |
| Foster (2019) | | | | | ✓ | | 17<br>(25 ± 4 y)<br>(10 m & 7 f) | 15<br>(75 ± 3 y)<br>(5 m & 10 f) | | | ✓ | 10 | Descent + ascent<br>4 steps | ✓ | Self-selected | |
| Hamel (2005) | | | ✓<br>Visual acuity | | ✓ | × | 12<br>(24.3 ± 2.5 y)<br>(All females) | 10<br>(73.5 ± 2.6 y)<br>(All females) | | | ✓ | | Descent + ascent<br>7 steps, each<br>Height: 18 cm<br>Width: 28 cm | ✓ | Fixed velocity<br>0.65 m/s ± 0.0325 m/s | |
| Hsue (2014) | | × | | | ✓ | | 28<br>(<40 y)<br>(12 m & 16 f) | | | 21<br>(>65 y)<br>(10 m & 11 f) | 2 | 8 | Descent + ascent<br>5 steps, each<br>Height: 18 cm<br>Width: 28 cm | ✓ | Self-selected | ✓ |
| Hsue (2009) | | | ✓ | | | | 16<br>(28.7 ± 5.6 y) | 10<br>(70.4 ± 4.4 y) | | | 2 | 8 | Descent + ascent<br>5 steps | ✓ | Self-selected | ✓ |

| | | | | | | | | | | | | | | | | |
|---|---|---|---|---|---|---|---|---|---|---|---|---|---|---|---|---|
| | | | Physical activity questionnaire | | | | (0 m & 16 f) | (0 m & 10 f) | | | | | | | | |
| Kim (2009) | ● | ✓ | ×<br>Berg Functional Balance Scale < 50<br>Frenchay Instrumental Activities of Daily Living score < 50<br>Physical Function score < 25<br>Visual acuity | ✓<br>Mini mental status | ✓ | | 15<br>(23.6 ± 2.4 y)<br>(5 m & 10 f) | | | 15<br>(73.1 ± 4.3 y)<br>(6 m & 9 f) | ✓ | | Descent + ascent<br>3 steps, each<br>Height: 17 cm<br>Width: 28 cm | ✓ | Self-selected | |
| Larsen (2008) | ● | | ×<br>Physical activity questionnaire | | ✓ | | 11<br>(25.8 ± 2 y) | | | 19<br>(72.3 ± 6.6 y) | 1 | | Descent + ascent<br>9 steps, each<br>Height: 16 cm<br>Width: 23 cm | ✓ | Self-selected | |
| Mian (2007) | ● | | ✓<br>Physical activity questionnaire<br>Short physical performance battery<br>Timed-Up-and-Go | ✓<br>Falls efficacy scale | ✓ | | 13<br>(28 ± 4 y)<br>(13 m & 0 f) | 15<br>(76 ± 3 y)<br>(15 m & 0 f) | | | ✓ | 9 | Descent + ascent<br>3 steps, each<br>Height: 16.5 cm<br>Width: 28 cm | ✓ | 90-95 steps/min | |
| Novak (2011) | ● | × | | | ✓ | | 24<br>(23.7 ± 3.0 y)<br>(7 m & 17 f) | | | 33<br>(67.0 ± 8.2 y)<br>(14 m & 19 f) | 1 | 2 | Descent + ascent<br>4 steps, each<br>Height: 15 cm<br>Width: 26 cm | ✓ | Self-selected | ✓ |
| Novak (2016) | ● | | | | ✓ | | 14<br>(25.5 ± 3.2 y)<br>(8 m & 6 f) | 14<br>(73.1 ± 6 y)<br>(7 m & 7 f) | | | | 3 | Descent + ascent<br>6 steps, each<br>Height: 7 to 8 inches<br>Width: 8 to 14 inches | × | Self-selected | ✓ |

| | | | | | | | | | | | | | | | | |
|---|---|---|---|---|---|---|---|---|---|---|---|---|---|---|---|---|
| Reeves (2009) | 🟠 | | | | | | 17<br>(24.6 ± 4.1 y)<br>(10 m & 7 f) | 15<br>(74.8 ± 2.8 y)<br>(5 m & 10 f) | | | 2 | 9 | Ascent<br>4 steps, each<br>Height: 17 cm<br>Width: 28 cm | ✓ | Self-selected | |
| **Perturbations** | | | | | | | | | | | | | | | | |
| Afschrift (2019) | 🟢 | ✓ | | | | | 18<br>(21 ± 2 y) | | | 10<br>(71 ± 4 y) | ✓ | 12 | Anteroposterior and mediolateral support translation | ✗ | 1.1 m/s | ✓ |
| Bosquée (2021) | 🟢 | | | | | | 12<br>(24 ± 3 y) | 11<br>(72 ± 5 y) | | | 1 | | Backward ankle pull | ✗ | Self-selected | ✓ |
| Debelle (2021) | 🟠 | | | | | | 17<br>(25.2 ± 3.7 y)<br>(8 m & 9 f) | 17<br>(62.4 ± 6.6 y)<br>(3 m & 14 f) | | | ✓ | 12 | Backward support acceleration at 5 $m/s^2$ | ✗ | Fixed velocity 1.2 m/s | ✓ |
| Jeon (2022a) | 🟠 | | | ✗<br>Mini mental status < 26 | | | 10<br>(28.2 ± 4.8 y)<br>(4 m & 6 f) | 10<br>(62.5 ± 4.7 y)<br>(7 m & 3 f) | | | | 8 | Anteroposterior support acceleration at 7.75 $m/s^2$, 12 $m/s^2$, 16.75 $m/s^2$ | ✗ | Self-selected | ✓ |
| Jeon (2022b) | 🟠 | | | | | | 10<br>(24 ± 3 y)<br>(4 m & 6 f) | | | 10<br>(77 ± 8 y)<br>(5 m & 5 f) | | 10 | 8 cm surface drop | ✗ | Self-selected | |
| Kazanski (2020) | 🟠 | | ✓<br>Timed-Up-and-Go<br>Square Step Test | ✓<br>Mini mental status examination<br>10-point abbreviated Iconographic-Falls Efficacy Scale | | | 17<br>(23.7 ± 3.7 y)<br>(8m & 9 f) | 17<br>(67.5 ± 4.9 y)<br>(7m & 10 f) | | | | 10 | Mediolateral support or visual field translation | ✓ | Self-selected | ✓ |
| Laudani (2021) | 🟠 | | ✗<br>Physical activity questionnaire (exclude highly active individuals)<br>✓<br>Berg Balance Scale | | ✓ | | 10<br>(25 ± 2 y)<br>(2 m & 8 f) | | 10<br>(73 ± 5 y)<br>(2 m & 8 f) | | 2 | 13 | Mediolateral waist-pull at 10% of the body mass | ✗ | Self-selected | ✓ |

| | | | | | | | | | | | | | | | | |
|---|---|---|---|---|---|---|---|---|---|---|---|---|---|---|---|---|
| Liu (2009) | 🟠 | | | | ✓ | | 9<br>(23.6 ± 4.8 y) | 9<br>(73.6 ± 4.4 y) | | | 2 | 6 | One of the force plates was covered in soapy water | × | | |
| Martelli (2017) | 🟠 | × | | | | × | 8<br>(24 ± 2.7 y) (4 m & 4 f) | | | 8<br>(65 ± 4.8 y)<br>(5 m & 3 f) | | 6 | Forward support acceleration at 0.89 $m/s^2$, 1.26 $m/s^2$, 1.54 $m/s^2$ | × | Fixed velocity<br>$V = \sqrt{Fr} * g * L$,<br>Fr = 0.15 | ✓ |
| McCrum (2016) | 🟠 | | | | | | 11<br>(25.5 ± 2.1 y)<br>(0 m & 11 f) | 14<br>(69.0 ± 4.7 y)<br>(0 m & 14 f) | | | | 8 | Backward ankle pull of 2.1 kg | × | 1.4 m/s | |
| McIntosh (2016) | 🟠 | | ✓<br>Berg Balance Scale | | ✓ | | 11<br>(23.8 ± 3.1 y)<br>(4 m & 7 f) | 10<br>(71.1 ± 3.1 y)<br>(2 m & 8 f) | | | | ✓ | Forward support translation of 16 cm at 130 $cm/s^2$ acceleration | × | Self-selected | ✓ |
| Nachmani (2020) | 🔴 | | | ✓<br>Mini mental status | ✓ | | 19<br>(26 ± 0.8 y)<br>No gender information | 35<br>(81 ± 4.5 y)<br>No gender information | | | | 2 | Medial-lateral support translation | × | Self-selected | ✓ |
| Qiao (2018) | 🟠 | × | ×<br>Health questionnaire<br>BMI > 30 | | | | 11<br>(24.8 ± 3.4 y)<br>(5 m & 6 f) | | | 11<br>(75.3 ± 5.4 y)<br>(5 m & 6 f) | | 14 | Mediolateral visual field translation | × | Self-selected | ✓ |
| Ren (2022) | 🟠 | × | ✓<br>Timed-Up-and-Go | | | | 15<br>(26.5 ± 3 y) | | | 15<br>(68.3 ± 3.3 y) | 2 | 10 | Anteroposterior support acceleration at 3 $m/s^2$ | × | Self-selected | ✓ |
| Roeles (2018) | 🟠 | | ✓<br>20 minute walking test | | ✓ | | 9<br>(25.1 ± 3.4 y)<br>(6 m & 3 f) | 9<br>(70.1 ± 8.1 y)<br>(2 m & 7 f) | | | | 32 | Mediolateral or anteroposterior support translations, or room darkening | × | Self-selected | ✓ |
| Rum (2020) | 🟠 | | ✓<br>Berg Balance Scale | ✓<br>Falls Efficacy Scale-International | ✓ | × | 10<br>(25 ± 2 y)<br>(2 m & 8 f) | 10<br>(73 ± 5y)<br>(2 m & 8 f) | | | 2 | 13 | Mediolateral waist-pull at 10% of the body mass | × | Self-selected | ✓ |

| | | | | | | | | | | | | | | | | |
|---|---|---|---|---|---|---|---|---|---|---|---|---|---|---|---|---|
| Shulman (2018) | | | ✓<br>Waterloo Footedness questionnaire | | ✓ | | 18<br>(21.7 ± 2.6 y)<br>(10 m & 8 f) | 16<br>(75.6 ± 5.3 y)<br>(9m & 7f) | | | 2 | 12 | Medial-lateral support translation at 60 cm/s, 2 m/s$^2$ | × | | ✓ |
| Shulman (2019) | | | | | ✓ | | 18<br>(21.7 ± 2.6 y)<br>(10 m & 8 f) | 16<br>(75.6 ± 5.3 y)<br>(9 m & 7 f) | | | 2 | 12 | Anterior-posterior support translation at 60 cm/s, 2 m/s$^2$ | × | | ✓ |
| Tropea (2015) | | | | | | | 6<br>(24 ± 1.7 y)<br>(4 m & 2 f) | 6<br>(66.7 ± 5.4 y)<br>(2 m & 4 f) | | | ✓ | ✓ | Forward support translation at 8 m/s$^2$ | × | Fixed velocity<br>V = $\sqrt{Fr} * g * L$,<br>Fr = 0.15 | |
| Yoo (2021) | | | | ✓<br>Montreal Cognitive Assessment | | | 14<br>(23.4 ± 2.9 y)<br>(7 m & 7 f) | 14<br>(70.9 ± 4.5 y)<br>(4 m & 10 f) | | | 2 | 12 | 10 m/s$^2$ forward support translation (split-belt treadmill) | × | Self-selected | ✓ |
| **Obstacles** | | | | | | | | | | | | | | | | |
| Caetano (2016) | | | ✓<br>Melbourne Edge Test<br>Physiological Performance Assessment<br>Trail Making Test | ✓<br>Montreal Cognitive Assessment<br>Iconographical-Fall Efficacy Scale | ✓ | | 21<br>(26 ± 4 y)<br>(9 m & 12 f) | | | 50<br>(74 ± 7 y)<br>(16 m & 34 f) | ✓ | | Visually projected obstacle<br>Height: 0<br>Length: between 1/3 of step length and 21.5 cm<br>Width: < 21.5 cm | ✓ | Self-selected | ✓ |
| Chen (1991) | | ✓ | ✓<br>Visual acuity, osteoarthritic symptoms, hearing loss, lower extremity weakness and reflexes, vibration sense | ✓<br>Mini mental status | ✓ | | 24<br>(21.7 ± 2.1 y)<br>(12 m & 12 f) | | | 24<br>(71.2 ± 5.5 y)<br>(12 m & 12 f) | | 4 | Height: 2.5, 5.1, 15.2 cm | ✓ | Self-selected | ✓ |
| Chen (1994) | | | ✓<br>Visual acuity, pain or numbness, hearing loss, | | ✓ | × | 24<br>(23.7 ± 2.1 y)<br>(12 m & 12 f) | 24<br>(73 ± 5.5 y)<br>(12 m & 12 f) | | | | ✓ | Visually projected obstacle | ✓ | Self-selected | ✓ |

| | | | | | | | | | | | | | | | | |
|---|---|---|---|---|---|---|---|---|---|---|---|---|---|---|---|---|
| | | | vibratory sense, lower extremity reflexes | | | | | | | | | | Height: 0<br>Length: 3 cm<br>Width: 70 cm | | | |
| Chien (2018) | 🟠 | × | | | ✓ | | 10<br>(28.1 ± 1 y)<br>(4 m & 6 f) | | | 10<br>(66.7 ± 5.21 y)<br>(3 m & 7 f) | | 8 | Height: 10 % of leg length<br>Length: 2 cm<br>Width: 60 cm | ✓ | Self-selected | ✓ |
| Draganich (2004) | 🟢 | | | | | | 10<br>(25.9 y)<br>(5 m & 5 f) | 10<br>(71.6 y)<br>(7 m & 3 f) | | | 1 | ✓ | Height: 20 cm<br>Length: 0.5 cm<br>Width: 90 cm | ✓ | Self-selected<br><0.85 m/s<br><1.2 m/s | ✓ |
| Eyal (2020) | 🟠 | × | ✓<br>Four step square test<br>Mini balance evaluation system<br>Timed-Up-and-Go | ✓<br>Montreal cognitive assessment<br>Color trails test | | | 20<br>(20-35 y)<br>(10 m & 10 f) | | | 20<br>(70-85 y)<br>(10 m & 10 f) | | | Height: 0.20, 0.5, 0.75, 1, 1.25 cm<br>Length: 20 cm<br>Width: 60 cm | × | Self-selected | ✓ |
| Hahn (2004) | 🟢 | | ✓<br>Berg Balance Scale | ✓<br>Mini mental status | ✓ | | 13<br>(25.7 ± 3.6 y)<br>(7 m & 6 f) | 13<br>(72.8 ± 6 y)<br>(8 m & 5 f) | | | | 6 | Height: 2.5 %, 5%, 10%, 15% of body height | ✓ | Self-selected | ✓ |
| Huang (2008) | 🟢 | | | | | | 10<br>(26.1 ± 2.5 y) | 15<br>(72 ± 6 y) | | | 2 | 7 | Height: 10 %, 20%, 30% of leg length | ✓ | Self-selected | |
| Kim (2013) | 🟢 | ✓ | ✓<br>Berg Balance Scale<br>Frenchay Activities Index<br>Physical Functioning score<br>Health Surveys | ✓<br>Mini mental status | ✓ | × | 9<br>(27 ± 3.6 y)<br>(4 m & 5 f) | | | 9<br>(75.1 ± 6.7y<br>(4 m & 5 f) | 2 | | Height: 10 cm | ✓ | Self-selected | ✓ |
| Kulkarni (2021) | 🟠 | | | | | | 17<br>(20.9 ± 1.9 y)<br>(3 m & 14 f) | 14<br>(69.7 ± 5.4y)<br>(4 m & 10 f) | | | | ✓ | Height: 23.5 cm<br>Length: 0.8 cm<br>Width: 100 cm | ✓ | Self-selected | |
| LoJacono (2018) | 🟠 | | ✓<br>Physical activity questionnaire | | | | 20<br>(22.5 ± 3.7 y) | 20<br>(55.6 ± 6 y) | | | | 12 | Height: 0.5 cm<br>Length: 10 cm<br>Width: 100 cm | ✓ | Self-selected | ✓ |
| Lowrey (2007) | 🟢 | | | ✓<br>Mini mental status | ✓ | × | 8<br>(23.1 ± 2 y)<br>(4 m & 4 f) | 8<br>(76.1 ± 4.3 y)<br>(4 m & 4 f) | | | | ✓ | Height: 45% of lower leg length | ✓ | Self-selected | ✓ |

| | | | | | | | | | | | | | | | | |
|---|---|---|---|---|---|---|---|---|---|---|---|---|---|---|---|---|
| | | | | | | | | | | | | | Length: 2.5 or 5 cm (unclear) | | | |
| Lu (2006) | | | | | | | 15<br>(23 ± 3 y)<br>No gender information | 15<br>(72 ± 6 y)<br>No gender information | | | 2 | 7 | Height: 10%, 20% and 30% of leg length | ✓ | Self-selected | |
| Luo (2022) | | × | | | ✓ | | 11<br>(27 ± 3 y)<br>(All males) | | | 7<br>(67.5 ± 2.5 y)<br>(All males) | | ✓ | Height: 14 cm | ✓ | Self-selected | ✓ |
| Maidan (2018) | | | ✓<br>Montreal cognitive assessment<br>Trail making test<br>Four square step test<br>Timed-Up-and-Go | ×<br>Mini mental status < 24 | ✓ | | 20<br>(29.3 ± 8.8 y)<br>(10 m & 10 f) | 20<br>(77.7 ± 3.5 y)<br>(10 m & 10 f) | | | | ✓ | Height: 2.5, 7.5 cm<br>Length: 20 cm<br>Width: 60 cm | × | Self-selected | ✓ |
| McFadyen (2002) | | | ✓<br>Foot vibration<br>Ostwestry back-pain questionnaire | | ✓ | | 10<br>(28.4 ± 5.4 y)<br>(All males) | 10<br>(69.5 ± 6.1 y)<br>(All males) | | | 2 | ✓ | Height: 5 cm<br>Length: 11.75 cm<br>Width: 122 cm | ✓ | Self-selected | ✓ |
| Mckenzie (2004) | | ✓ | ✓<br>Standard sensorimotor tests | ✓<br>Mini mental status | | | 15<br>(22.5 ± 2.77 y)<br>(5 m & 10 f) | | | 17<br>(68.94 ± 4.85 y)<br>(7 m & 10 f) | | 6 | Height: 15 cm<br>Width: 60, 15 cm | ✓ | Self-selected | ✓ |
| Park (2012) | | | ✓<br>Berg Balance Scale | ✓<br>Mini mental status | | | 9<br>(25.8 ± 2.8 y)<br>(3 m & 6 f) | 9<br>(69.7 ± 3.9 y)<br>(3 m & 6 f) | | | | 6 | Height: 10%, 20%, 30% of leg length<br>Width: 100 cm | ✓ | Self-selected | ✓ |
| Tomar (2012) | | | ✓<br>Berg Balance Scale<br>Trail-making test<br>Timed-Up-and-Go<br>Choice stepping reaction time | ✓<br>Mini mental status | | | 30<br>(18-30 y)<br>(All males) | 30<br>(>65 y)<br>(All males) | | | | 1 | Height: 7.6, 12.7, 20.3 cm<br>Length: 12.7 cm<br>Width: 25.4 cm | ✓ | Self-selected | |
| Uchiyama (2012) | | | ✓<br>Berg Balance Scale | | | | 17<br>(21 ± 2.4 y) | 30<br>(70 ± 6.9 y) | | | | | Height: 5 cm<br>Length: 10 cm<br>Width: 120 cm | ✓ | Self-selected | ✓ |

| | | | | | | | | | | | | | | | | |
|---|---|---|---|---|---|---|---|---|---|---|---|---|---|---|---|---|
| Wang (2010) | ● | | ✓<br>Snellen vision test | | | × | 15<br>(23 ± 3 y)<br>(8 m & 7 f) | 15<br>(72 ± 6 y)<br>(8 m & 7 f) | | | 2 | 7 | Height: 10%, 20% and 30% of leg length | ✓ | Self-selected | |
| Weerdesteyn (2005a) | ● | ✓ | | | | | 25<br>(20-37 y)<br>(4 m & 21 f) | | 99<br>(65-88 y)<br>(22 m & 77 f)<br>Only subjects with fall history were included | | | ✓ | Height: 1.5 cm<br>Length: 30 cm<br>Width: 40 cm | × | Fixed velocity 0.83 m/s | ✓ |
| Weerdesteyn (2005b) | ● | | Recruited from elderly sports groups | | | | 10<br>(19-32 y)<br>(0 m & 10 f) | | | 10<br>(65-78 y)<br>(0 m & 10 f) | | 6 | Height: 1.5 cm<br>Length: 30 cm<br>Width: 40 cm | × | Self-selected | ✓ |
