## Appendix E - Ageing references for "Systematic review of candidate prognostic factors for falling in older adults identified from motion analysis of challenging walking tasks"

**Appendix E – References of ageing studies**


Afschrift, M., van Deursen, R., De Groote, F., & Jonkers, I. (2019). Increased use of stepping strategy in response to medio-lateral perturbations in the elderly relates to altered reactive tibialis anterior activity. Gait & Posture, 68, 575-582. https://doi.org/10.1016/j.gaitpost.2019.01.010

Begg, R. K., & Sparrow, W. A. (2000). Gait characteristics of young and older individuals negotiating a raised surface : Implications for the prevention of falls. Journals of Gerontology - Series A Biological Sciences and Medical Sciences, 55(3), 147-154. https://doi.org/10.1093/gerona/55.3.M147

Bosquée, J., Werth, J., Epro, G., Hülsdünker, T., Potthast, W., Meijer, K., Ellegast, R., & Karamanidis, K. (2021). The ability to increase the base of support and recover stability is limited in its generalisation for different balance perturbation tasks. European Review of Aging and Physical Activity, 18(1), 1-10. https://doi.org/10.1186/s11556-021-00274-w

Bosse, I., Oberländer, K. D., Savelberg, H. H., Meijer, K., Brüggemann, G. P., & Karamanidis, K. (2012). Dynamic stability control in younger and older adults during stair descent. Human Movement Science, 31(6), 1560-1570. https://doi.org/10.1016/j.humov.2012.05.003

Caetano, M. J. D., Lord, S. R., Schoene, D., Pelicioni, P. H. S., Sturnieks, D. L., & Menant, J. C. (2016). Age-related changes in gait adaptability in response to unpredictable obstacles and stepping targets. Gait and Posture, 46, 35-41. https://doi.org/10.1016/j.gaitpost.2016.02.003

Chen, H. C., Ashton-Miller, J. A., Alexander, N. B., & Schultz, A. B. (1991). Stepping over obstacles : Gait patterns of healthy young and old adults. Journals of Gerontology, 46(6), 196-203. https://doi.org/10.1093/geronj/46.6.M196

Chen, H. C., Ashton-Miller, J. A., Alexander, N. B., & Schultz, A. B. (1994). Effects of age and available response time on ability to step over an obstacle. Journal of Gerontology, 49(5), M227-233. https://doi.org/10.1093/geronj/49.5.m227

Chien, J. H., Post, J., & Siu, K. C. (2018). Effects of Aging on the Obstacle Negotiation Strategy while Stepping over Multiple Obstacles. Scientific Reports, 8(1), 1-9. https://doi.org/10.1038/s41598-018-26807-5

Chiu, S. L., Chang, C. C., Dennerlein, J. T., & Xu, X. (2015). Age-related differences in inter-joint coordination during stair walking transitions. Gait and Posture, 42(2), 152-157. https://doi.org/10.1016/j.gaitpost.2015.05.003

Christina, K. A., & Cavanagh, P. R. (2002). Ground reaction forces and frictional demands during stair descent : Effects of age and illumination. Gait and Posture, 15(2), 153-158. https://doi.org/10.1016/S0966-6362(01)00164-3

Crosbie, J., & Gan, N. (2003). Effect of age and visual contrast on gait during obstacle negotiation. Australasian Journal on Ageing, 22(3), 131-135. https://doi.org/10.1111/j.1741-6612.2003.tb00483.x

Debelle, H., Maganaris, C. N., & O'Brien, T. D. (2021). Biomechanical Mechanisms of Improved Balance Recovery to Repeated Backward Slips Simulated by Treadmill Belt Accelerations in Young and Older Adults. Frontiers in Sports and Active Living, 3(September), 1-15. https://doi.org/10.3389/fspor.2021.708929

Dewolf, A. H., Sylos-Labini, F., Cappellini, G., Zhvansky, D., Willems, P. A., Ivanenko, Y., & Lacquaniti, F. (2021). Neuromuscular Age-Related Adjustment of Gait When Moving Upwards and Downwards. Frontiers in Human Neuroscience, 15(October), 1-14. https://doi.org/10.3389/fnhum.2021.749366

Dixon, P. C., Stirling, L., Xu, X., Chang, C. C., Dennerlein, J. T., & Schiffman, J. M. (2018). Aging may negatively impact movement smoothness during stair negotiation. Human Movement Science, 60, 78-86. https://doi.org/10.1016/j.humov.2018.05.008

Draganich, L. F., & Kuo, C. E. (2004). The effects of walking speed on obstacle crossing in healthy young and healthy older adults. Journal of Biomechanics, 37(6), 889-896. https://doi.org/10.1016/j.jbiomech.2003.11.002

Eyal, S., Kurz, I., Mirelman, A., Maidan, I., Giladi, N., & Hausdorff, J. M. (2020). Successful Negotiation of Anticipated and Unanticipated Obstacles in Young and Older Adults : Not All Is as Expected. Gerontology, 66(2), 187-196. https://doi.org/10.1159/000502140

Foster, R. J., Maganaris, C. N., Reeves, N. D., & Buckley, J. G. (2019). Centre of mass control is reduced in older people when descending stairs at an increased riser height. Gait & Posture, 73, 305-314. https://doi.org/10.1016/j.gaitpost.2019.08.004

Francksen, N., Ackermans, T., Holzer, D., Maganaris, C., Hollands, M., Roys, M., & O'Brien, T. (2022). Underlying mechanisms of fall risk on stairs with inconsistent going size. Applied ergonomics, 101, e103678-e103678. https://doi.org/10.1016/j.apergo.2022.103678

Francksen, N. C., Ackermans, T. M. A., Holzer, D., Ebner, S. A., Maganaris, C. N., Hollands, M. A., Karamanidis, K., Roys, M., & O'Brien, T. D. (2020). Negotiating stairs with an inconsistent riser : Implications for stepping safety. Applied Ergonomics, 87(July 2019), 103131. https://doi.org/10.1016/j.apergo.2020.103131

Hahn, M. E., & Chou, L.-S. (2004). Age-related reduction in sagittal plane center of mass motion during obstacle crossing. Journal of Biomechanics, 37(6), 837-844. https://doi.org/10.1016/j.jbiomech.2003.11.010

Hamel, K. A., Okita, N., Bus, S. A., & Cavanagh, P. R. (2005). A comparisson of foot/ground interaction during stair negotiation and level walking in young and older women. Ergonomics, 48(8), 1047-1056. https://doi.org/10.1080/00140130500193665

Hsue, B. J., & Su, F. C. (2009). Kinematics and kinetics of the lower extremities of young and elder women during stairs ascent while wearing low and high-heeled shoes. Journal of Electromyography and Kinesiology, 19(6), 1071-1078. https://doi.org/10.1016/j.jelekin.2008.09.005

Hsue, B.-J., & Su, F.-C. (2014). Effects of Age and Gender on Dynamic Stability During Stair Descent. Archives of Physical Medicine and Rehabilitation, 95(10), 1860-1869. https://doi.org/10.1016/j.apmr.2014.05.001

Huang, S. C., Lu, T. W., Chen, H. L., Wang, T. M., & Chou, L. S. (2008). Age and height effects on the center of mass and center of pressure inclination angles during obstacle-crossing. Medical Engineering and Physics, 30(8), 968-975. https://doi.org/10.1016/j.medengphy.2007.12.005

Jeon, W., Wang, S., Bhatt, T., & Westlake, K. P. (2022a). Perturbation-Induced Protective Arm Responses : Effect of Age, Perturbation-Intensity, and Relationship with Stepping Stability : A Pilot Study. Brain Sciences, 12(7). https://doi.org/10.3390/brainsci12070953

Jeon, W., Whitall, J., & Westlake, K. (2022b). Age-related differences in stepping stability following a sudden gait perturbation are associated with lower limb eccentric control of the perturbed limb. Experimental Gerontology, 167(August), 111917. https://doi.org/10.1016/j.exger.2022.111917

Kazanski, M. E., Cusumano, J. P., & Dingwell, J. B. (2020). How healthy older adults regulate lateral foot placement while walking in laterally destabilizing environments. Journal of Biomechanics, 104, 109714. https://doi.org/10.1016/j.jbiomech.2020.109714

Kim, H. D. (2009). A comparison of the center of pressure during stair descent in young and healthy elderly adults. Journal of Physical Therapy Science, 21(2), 129-134. https://doi.org/10.1589/jpts.21.129

Kim, K.-M., Hart, J. M., & Hertel, J. (2013). Influence of body position on fibularis longus and soleus Hoffmann reflexes. Gait & Posture, 37(1), 138-140. https://doi.org/10.1016/j.gaitpost.2012.06.009

Kulkarni, A., Cho, H. Y., Rietdyk, S., & Ambike, S. (2021). Step length synergy is weaker in older adults during obstacle crossing. Journal of Biomechanics, 118, 110311. https://doi.org/10.1016/j.jbiomech.2021.110311

Larsen, A. H., Puggaard, L., Hämäläinen, U., & Aagaard, P. (2008). Comparison of ground reaction forces and antagonist muscle coactivation during stair walking with ageing. Journal of Electromyography and Kinesiology, 18(4), 568-580. https://doi.org/10.1016/j.jelekin.2006.12.008

Laudani, L., Rum, L., Valle, M. S., Macaluso, A., Vannozzi, G., & Casabona, A. (2021). Age differences in anticipatory and executory mechanisms of gait initiation following unexpected balance perturbations. European Journal of Applied Physiology, 121(2), 465-478. https://doi.org/10.1007/s00421-020-04531-1

Liu, J., & Lockhart, T. E. (2009). Age-related joint moment characteristics during normal gait and successful reactive-recovery from unexpected slip perturbations. Gait & Posture, 30(3), 276-281. https://doi.org/10.1016/j.gaitpost.2009.04.005

Lojacono, B. C. T., Macpherson, R. P., Kuznetsov, N. A., Raisbeck, L. D., Ross, E., Rhea, C. K., & Raisbeck, L. D. (2018). Journal of Motor Learning and Development, 6 (2), 234-249.

Lowrey, C. R., Watson, A., & Vallis, L. A. (2007). Age-related changes in avoidance strategies when negotiating single and multiple obstacles. Experimental Brain Research, 182(3), 289-299. https://doi.org/10.1007/s00221-007-0986-0

Lu, T. W., Chen, H. L., & Chen, S. C. (2006). Comparisons of the lower limb kinematics between young and older adults when crossing obstacles of different heights. Gait and Posture, 23(4), 471-479. https://doi.org/10.1016/j.gaitpost.2005.06.005

Luo, Y., Yang, F., Yerebakan, M. O., Zhang, J., & Hu, B. (2022). Load Carriage Modes and Limb Crossing Patterns Altered Gait during Obstacle Negotiation. Journal of Motor Behavior, 54(5), 525-536. https://doi.org/10.1080/00222895.2021.2017837

Maidan, I., Eyal, S., Kurz, I., Geffen, N., Gazit, E., Ravid, L., Giladi, N., Mirelman, A., & Hausdorff, J. M. (2018). Age-associated changes in obstacle negotiation strategies : Does size and timing matter? Gait & Posture, 59, 242-247. https://doi.org/10.1016/j.gaitpost.2017.10.023

Martelli, D., Aprigliano, F., Tropea, P., Pasquini, G., Micera, S., & Monaco, V. (2017). Stability against backward balance loss : Age-related modifications following slip-like perturbations of multiple amplitudes. Gait and Posture, 53, 207-214. https://doi.org/10.1016/j.gaitpost.2017.02.002

McCrum, C., Epro, G., Meijer, K., Zijlstra, W., Brüggemann, G.-P., & Karamanidis, K. (2016). Locomotor stability and adaptation during perturbed walking across the adult female lifespan. Journal of Biomechanics, 49(7), 1244-1247. https://doi.org/10.1016/j.jbiomech.2016.02.051

McFadyen, B. J., & Prince, F. (2002). Avoidance and accommodation of surface height changes by healthy, community-dwelling, young, and elderly men. Journals of Gerontology - Series A Biological Sciences and Medical Sciences, 57(4), B166-B174. https://doi.org/10.1093/gerona/57.4.B166

McIntosh, E. I., Zettel, J. L., & Vallis, L. A. (2017). Stepping Responses in Young and Older Adults Following a Perturbation to the Support Surface During Gait. Journal of Motor Behavior, 49(3), 288-298. https://doi.org/10.1080/00222895.2016.1204262

McKenzie, N. C., & Brown, L. A. (2004). Obstacle negotiation kinematics : Age-dependent effects of postural threat. Gait and Posture, 19(3), 226-234. https://doi.org/10.1016/S0966-6362(03)00060-2

Mian, O. S., Narici, M. V., Minetti, A. E., & Baltzopoulos, V. (2007). Centre of mass motion during stair negotiation in young and older men. Gait & Posture, 26(3), 463-469. https://doi.org/10.1016/j.gaitpost.2006.11.202

Nachmani, H., Shani, G., Shapiro, A., & Melzer, I. (2020). Characteristics of first recovery step response following unexpected loss of balance during walking : A dynamic approach. Gerontology, 66(4), 362-370. https://doi.org/10.1159/000505649

Novak, A. C., & Brouwer, B. (2011). Sagittal and frontal lower limb joint moments during stair ascent and descent in young and older adults. Gait and Posture, 33(1), 54-60. https://doi.org/10.1016/j.gaitpost.2010.09.024

Novak, A. C., Komisar, V., Maki, B. E., & Fernie, G. R. (2016). Age-related differences in dynamic balance control during stair descent and effect of varying step geometry. Applied Ergonomics, 52, 275-284. https://doi.org/10.1016/j.apergo.2015.07.027

Park, S. Y., & Lee, Y. S. (2012). Kinematics of the lower limbs during obstacle crossings performed by young adults and the elderly. Journal of Physical Therapy Science, 24(10), 941-944. https://doi.org/10.1589/jpts.24.941

Qiao, M., Feld, J. A., & Franz, J. R. (2018). Aging effects on leg joint variability during walking with balance perturbations. Gait & Posture, 62, 27-33. https://doi.org/10.1016/j.gaitpost.2018.02.020

Reeves, N. D., Spanjaard, M., Mohagheghi, A. A., Baltzopoulos, V., & Maganaris, C. N. (2009). Older adults employ alternative strategies to operate within their maximum capabilities when ascending stairs. Journal of Electromyography and Kinesiology, 19(2). https://doi.org/10.1016/j.jelekin.2007.09.009

Ren, X., Lutter, C., Kebbach, M., Bruhn, S., Bader, R., & Tischer, T. (2022). Lower extremity joint compensatory effects during the first recovery step following slipping and stumbling perturbations in young and older subjects. BMC Geriatrics, 22(1), 1-16. https://doi.org/10.1186/s12877-022-03354-3

Roeles, S., Rowe, P. J., Bruijn, S. M., Childs, C. R., Tarfali, G. D., Steenbrink, F., & Pijnappels, M. (2018). Gait stability in response to platform, belt, and sensory perturbations in young and older adults. Medical and Biological Engineering and Computing, 56(12), 2325-2335. https://doi.org/10.1007/s11517-018-1855-7

Rum, L., Vannozzi, G., Macaluso, A., & Laudani, L. (2021). Neuromechanical response of the upper body to unexpected perturbations during gait initiation in young and older adults. Aging Clinical and Experimental Research, 33(4), 909-919. https://doi.org/10.1007/s40520-020-01592-2

Shulman, D., Spencer, A., & Ann Vallis, L. (2019). Older adults exhibit variable responses in stepping behaviour following unexpected forward perturbations during gait initiation. Human Movement Science, 63(May 2018), 120-128. https://doi.org/10.1016/j.humov.2018.11.008

Shulman, D., Spencer, A., & Vallis, L. A. (2018). Age-related alterations in reactive stepping following unexpected mediolateral perturbations during gait initiation. Gait and Posture, 64(August 2017), 130-134. https://doi.org/10.1016/j.gaitpost.2018.05.035

Tomar, U. S., & Gupta, N. (2012). An observational study of foot lifts asymmetry during obstacle avoidance. Journal of Neurosciences in Rural Practice, 3(3), 324-327. https://doi.org/10.4103/0976-3147.102614

Tropea, P., Martelli, D., Aprigliano, F., Micera, S., & Monaco, V. (2015). Effects of aging and perturbation intensities on temporal parameters during slipping-like perturbations. Proceedings of the Annual International Conference of the IEEE Engineering in Medicine and Biology Society, EMBS, 2015-Novem, 5291-5294. https://doi.org/10.1109/EMBC.2015.7319585

Uchiyama, M., Demura, S., & Sugiura, H. (2012). The mobility performance of the elderly before, during and after crossing over an obstacle. Human Movement, 13(4), 297-302. https://doi.org/10.2478/v10038-012-0034-1

Wang, T. M., Chen, H. L., Hsu, W. C., Liu, M. W., & Lu, T. W. (2010). Biomechanical role of the locomotor system in controlling body center of mass motion in older adults during obstructed Gait. Journal of Mechanics, 26(2), 195-203. https://doi.org/10.1017/S1727719100003051

Weerdesteyn, V., Nienhuis, B., & Duysens, J. (2005a). Advancing age progressively affects obstacle avoidance skills in the elderly. Human Movement Science, 24(5-6), 865-880. https://doi.org/10.1016/j.humov.2005.10.013

Weerdesteyn, V., Nienhuis, B., Mulder, T., & Duysens, J. (2005b). Older women strongly prefer stride lengthening to shortening in avoiding obstacles. Experimental Brain Research, 161(1), 39-46. https://doi.org/10.1007/s00221-004-2043-6

Yoo, D., An, J., Seo, K. H., & Lee, B. C. (2021). Aging Affects Lower Limb Joint Moments and Muscle Responses to a Split-Belt Treadmill Perturbation. Frontiers in Sports and Active Living, 3(July), 1-12. https://doi.org/10.3389/fspor.2021.683039
