## Appendix F - Ageing outcomes for "Systematic review of candidate prognostic factors for falling in older adults identified from motion analysis of challenging walking tasks"

**Appendix F**: Significant and non-significant findings in ageing studies
Overview of the reported outcome parameters and corresponding articles reporting either significant or non-significant findings. Studies with a serious risk of bias are in red.
Sign., significant; N.S. non-significant.

| **Success and error** | **Total** | | **Stairs** | | **Perturbations** | | **Obstacles** | |
|---|---|---|---|---|---|---|---|---|
| | Sign. | N.S. | Sign. | N.S. | Sign. | N.S. | Sign. | N.S. |
| Stepping error, accuracy, and success | 6 | 3 | | | Nachmani (2020) | Debelle (2021) | Caetano (2016)<br>Chen (1994)<br>Kim (2013)<br>Weerdesteyn (2005a)<br>Weerdesteyn (2005b) | Eyal (2020)<br>LoJacono (2018) |
| **Temporal outcomes** | **Total** | | **Stairs** | | **Perturbations** | | **Obstacles** | |
| | Sign. | N.S. | Sign. | N.S. | Sign. | N.S. | Sign. | N.S. |
| Reaction time | 0 | 1 | | | | | | Weerdesteyn (2005a) |
| Cadence | 1 | 2 | | | | Martelli (2017) | Caetano (2016) | McFadyen (2002) |
| **Foot and head outcomes** | **Total** | | **Stairs** | | **Perturbations** | | **Obstacles** | |
| | Sign. | N.S. | Sign. | N.S. | Sign. | N.S. | Sign. | N.S. |
| Foot velocity | 0 | 1 | | | | | | Luo (2022) |
| Foot velocity variation | 0 | 1 | | | | | | Luo (2022) |
| Head velocity | 1 | 0 | Dixon (2018) | | | | | |
| Power spectrum head velocity | 1 | 0 | Dixon (2018) | | | | | |
| **Center of Mass (CoM) outcomes** | **Total** | | **Stairs** | | **Perturbations** | | **Obstacles** | |
| | Sign. | N.S. | Sign. | N.S. | Sign. | N.S. | Sign. | N.S. |
| Power spectrum CoM velocity | 1 | 0 | Dixon (2018) | | | | | |
| CoM trajectory and jerk score | 3 | 2 | Dixon (2018) | Bosse (2012) | Laudani (2021) | | Wang (2010) | Lowrey (2007) |
| Extrapolated CoM | 2 | 1 | Bosse (2012) | Novak (2016) | Afschrift (2019) | | | |
| **Force plate outcomes** | **Total** | | **Stairs** | | **Perturbations** | | **Obstacles** | |
| | Sign. | N.S. | Sign. | N.S. | Sign. | N.S. | Sign. | N.S. |
| CoP displacement and velocity | 3 | 1 | Kim (2009) | | Afschrift (2019)<br>Jeon (2022b) | Rum (2020) | | |
| Base of Support | 1 | 1 | | | Bosquée (2021) | McCrum (2016) | | |
| Support moment peak and variability | 1 | 0 | Novak (2011) | | | | | |
| **Dynamic stability outcomes** | **Total** | | **Stairs** | | **Perturbations** | | **Obstacles** | |
| | Sign. | N.S. | Sign. | N.S. | Sign. | N.S. | Sign. | N.S. |
| Angle between CoM and CoP | 1 | 0 | Huang (2008) | | | | | |
| Distance between CoM and BoS | 1 | 0 | | | Nachmani (2020) | | | |

| | | | | | | | | | |
|---|---|---|---|---|---|---|---|---|---|
| Relative MoS to baseline | | 1 | 0 | | | McCrum (2016) | | | |
| CoM-CoP distance | | 5 | 1 | Hsue (2014)<br>Huang (2008)<br>Reeves (2009) | Mian (2007) | | | Hahn (2004)<br>Wang (2010) | |
| CoM-CoP angular velocity | | 0 | 1 | | Huang (2008) | | | | |
| **Joint and segment outcomes** | | **Total** | | **Stairs** | | **Perturbations** | | **Obstacles** | |
| | | Sign. | N.S. | Sign. | N.S. | Sign. | N.S. | Sign. | N.S. |
| Ankle | motion | 5 | 3 | Hsue (2009) | Dewolf (2021)<br>Reeves (2009) | | | Chen (1991)<br>Lu (2006)<br>McFadyen (2002)<br>Park (2012) | Draganich (2004) |
| | motion variability | 1 | 0 | | | Qiao (2018) | | | |
| | moment rate | 1 | 0 | | | Liu (2009) | | | |
| | power | 1 | 0 | | | | | McFadyen (2002) | |
| | work | 0 | 1 | | Foster (2019) | | | | |
| | angular impulse | 1 | 0 | Bosse (2012) | | | | | |
| Knee | motion | 5 | 5 | Bosse (2012)<br>Hsue (2009) | Reeves (2009) | Jeon (2022b) | | Chien (2018)<br>Park (2012) | Chen (1991)<br>Draganich (2004)<br>Lu (2006)<br>McFadyen (2002) |
| | motion variability | 1 | 0 | | | Qiao (2018) | | | |
| | moment rate | 1 | 0 | | | Liu (2009) | | | |
| | power | 0 | 1 | | | | Jeon (2022b) | | |
| | work | 1 | 1 | | Foster (2019) | Jeon (2022b) | | | |
| | angular impulse | 1 | 0 | Bosse (2012) | | | | | |
| | power | 1 | 0 | | | | | McFadyen (2002) | |
| Hip | motion variability | 1 | 0 | | | Qiao (2018) | | | |
| | moment rate | 1 | 0 | | | Liu (2009) | | | |
| | flexion velocity | 1 | 0 | | | | | Draganich (2004) | |
| Ankle, knee and hip coordination patterns | | 1 | 0 | Chiu (2015) | | | | | |
| Shank | Range of Motion | 1 | 0 | Dewolf (2021) | | | | | |
| | elevation angle | 0 | 1 | | Dewolf (2021) | | | | |
| Thigh | Range of Motion | 1 | 0 | Dewolf (2021) | | | | | |
| | elevation angle | 0 | 1 | | Dewolf (2021) | | | | |
| Thigh-shank phase lag | | 1 | 0 | Dewolf (2021) | | | | | |
| Arm | displacement | 0 | 1 | | | | Jeon (2022a) | | |
| | displacement velocity | 0 | 1 | | | Jeon (2022a) | | | |
| Body tilt | | 2 | 4 | Novak (2016) | Bosse (2012)<br>Hsue (2009) | Rum (2020) | Debelle (2021)<br>Martelli (2017) | | |
